# Determinants of anti-PD1 response and resistance in clear cell renal cell carcinoma

**DOI:** 10.1101/2021.03.19.21253661

**Authors:** Lewis Au, Emine Hatipoglu, Marc Robert de Massy, Kevin Litchfield, Andrew Rowan, Rachael Thompson, Desiree Schnidrig, Fiona Byrne, Gordon Beattie, Stuart Horswell, Nicos Fotiadis, Steve Hazell, David Nicol, Scott Thomas Colville Shepherd, Annika Fendler, Robert Mason, Jan Attig, Kroopa Joshi, Imran Uddin, Pablo Becker, Mariana Werner Sunderland, Ayse Akarca, Ignazio Puccio, William Yang, Tom Lund, Kim Dhillon, Marcos Duran Vasquez, Ehsan Ghorani, Hang Xu, José Ignacio López, Anna Green, Ula Mahadeva, Elaine Borg, Miriam Mitchison, David Moore, Ian Proctor, Mary Falzon, Andrew Furness, Lisa Pickering, James L. Reading, Roberto Salgado, Teresa Marafioti, Mariam Jamal-Hanjani, on behalf of the PEACE Consortium, George Kassiotis, Benny Chain, James Larkin, Charles Swanton, Sergio A Quezada, Samra Turajlic, on behalf of the TRACERx Renal Consortium

**Affiliations:** Cancer Dynamics Laboratory, The Francis Crick Institute, London NW1 1AT, UK; Renal and Skin Units, The Royal Marsden NHS Foundation Trust, London SW3 6JJ, UK; Cancer Immunology Unit, Research Department of Haematology, University College London Cancer Institute, London WC1E 6DD, UK; Cancer Research UK Lung Cancer Centre of Excellence, University College London Cancer Institute, London WC1E 6DD, UK; Cancer Evolution and Genome Instability Laboratory, The Francis Crick Institute, London NW1 1AT, UK; Retroviral Immunology, The Francis Crick Institute, London NW1 1AT, UK; Department of Bioinformatics and Biostatistics, The Francis Crick Institute, London NW1 1AT, UK; Cancer Research UK Cancer Imaging Centre, Division of Radiotherapy and Imaging, The Institute of Cancer Research and Royal Marsden Hospital, London SW3 6JJ, UK; Department of Pathology, the Royal Marsden NHS Foundation Trust, London SW3 6JJ, UK; Department of Urology, the Royal Marsden NHS Foundation Trust, London SW3 6JJ, UK; Department of Cellular Pathology, University College London Hospital, London NW1 2BU, UK; Department of Pathology, Cruces University Hospital, Biocruces-Bizkaia Institute, 48903 Barakaldo, Bizkaia, Spain; Department of Cellular Pathology, Guy’s & St Thomas’ NHS Foundation Trust, St Thomas’ Hospital, London SE1 7EH, UK; Division of Research, Peter MacCallum Cancer Centre, Melbourne VIC300, Australia; Department of Pathology, GZA-ZNA Hospitals, Wilrijk, Antwerp, Belgium; Cancer Metastasis Laboratory, University College London Cancer Institute, London WC1E 6DD, UK; Department of Medical Oncology, University College London Hospitals, London NW1 2BU, UK; Division of Infection and Immunity, University College London, London WC1E 6BT, UK; University College London Cancer Institute, London WC1E 6DD, UK

**Keywords:** clear cell renal cell carcinoma, nivolumab, immunotherapy, cytotoxicity, TCR clonal maintenance, TCR clonal replacement, human endogenous retrovirus, longitudinal sampling, multiregion sampling, autopsy

## Abstract

Antigen recognition and T-cell mediated cytotoxicity in clear-cell renal cell carcinoma (ccRCC) remains incompletely understood. To address this knowledge gap, we analysed 115 multiregion tumour samples collected from 15 treatment-naïve patients pre- and post-nivolumab therapy, and at autopsy in three patients. We performed whole-exome sequencing, RNAseq, TCRseq, multiplex immunofluorescence and flow cytometry analyses and correlated with clinical response. We observed pre-treatment intratumoural TCR clonal expansions suggesting pre-existing immunity. Nivolumab maintained pre-treatment expanded, clustered TCR clones in responders, suggesting ongoing antigen-driven stimulation of T-cells. T-cells in responders were enriched for expanded TCF7^+^CD8^+^ T-cells and upregulated GZMK/B upon nivolumab-binding. By contrast, nivolumab promoted accumulation of new TCR clones in non-responders, replacing pre-treatment expanded clonotypes. In this dataset, mutational features did not correlate with response to nivolumab and human endogenous retrovirus expression correlated indirectly. Our data suggests that nivolumab potentiates clinical responses in ccRCC by binding pre-existing expanded CD8^+^ T-cells to enhance cytotoxicity.

## Introduction

Clear-cell renal cell carcinoma (ccRCC) is the most common histological subtype of kidney cancer^1^ with a rising global incidence^2^. Instances of spontaneous regression^3-5^, efficacy of interleukin-2^6, 7^ and immune checkpoint inhibitors (CPI)^8-11^ confirm ccRCC as an immunogenic tumour type, but the nature of the antigenic stimulus is unknown. ccRCC carries a modest tumour mutational burden (TMB) (median of 1.42 mutations per megabase (mut/mb))^12^, ten-fold lower than melanoma and comparable to immune ‘cold’ tumours^13^. In contrast to melanoma^14^, non-small cell lung cancer^15, 16^, bladder^17^, and colorectal cancers^18^, TMB does not associate with CPI response in ccRCC^19-21^. Kidney tumours are enriched for frameshift insertion and deletions (fsINDELs)^22^, which can generate novel open-reading frames triggering a large number of highly distinct neoantigens. However, fsINDEL burden has so far not been shown to predict benefit from CPI in patients with ccRCC^20, 21^, again in contrast to other tumour types^22, 23^. Mutations in *PBRM1* are reported to be enriched in responders to CPI in ccRCC^19, 24, 25^, though this has not been observed consistently^20, 21, 26, 27^.

Large-scale, pan-cancer transcriptional analyses have shown ccRCCs is amongst the most highly immune-infiltrated solid tumour types^1, 28^. However, in contrast to other cancers, immune infiltration correlates with poorer prognosis in ccRCC^29^. The baseline composition of the infiltrate has been linked to CPI benefit, with high T-cell/low myeloid infiltration and high B-cell abundance enriched in responders to atezolizumab (anti-PDL1)^20^ and nivolumab (anti-PD1)^30^, respectively.

Intratumour heterogeneity (ITH) can impact therapy response in cancer through clonal selection of resistance mechanisms, loss of HLA heterozygosity, and loss of clonal neoantigens^31-33^. ITH is a frequent feature of ccRCC that associates with outcomes following surgery, but its impact on therapy response is unknown^34-36^. Furthermore, ITH complicates evaluation of prognostic and predictive biomarkers in all settings.

ADAPTeR (NCT02446860) is a phase II, single-arm, open-label study of nivolumab in treatment-naive metastatic ccRCC. Patients underwent multiregional fresh tumour sampling of primary and/or metastatic sites at baseline, week-9, at surgery (if performed), and disease progression. Key aim of the study was to evaluate genomic and tumour immune microenvironment features, throughout therapy. Patients were co-recruited to TRACERx Renal (TRAcking Cancer Evolution through therapy[Rx]; NCT03226886), and the PEACE (Posthumous Evaluation of Advanced Cancer Environment; NCT03004755) studies to expand the spatial and temporal breadth of profiling. We present an integrated analysis of clinical features and whole-exome and RNA sequencing, TCR profiling and multiplex immunohistochemistry/immunofluorescence (mIHC/IF); and high dimensional flow cytometry across longitudinal, multiregion fresh tumour samples in this cohort (**Figure 1A**).

**Figure 1.**
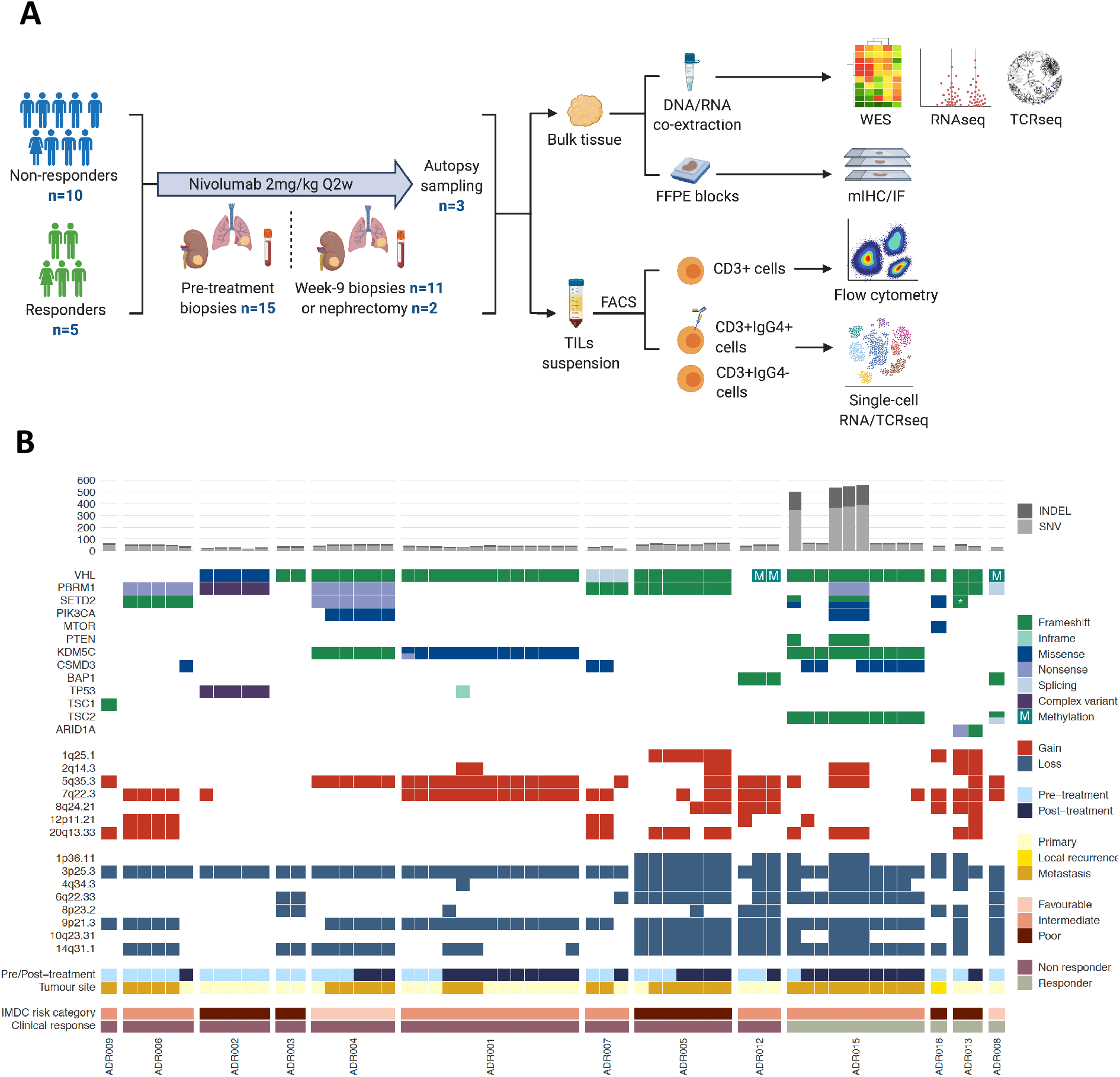
Experimental workflow, patients and samples overview, and genomic characteristics of the ADAPTeR cohort. **(A)** Overview of experimental workflow. Number (n) of patients contributing to sample collection at different timepoints are shown. **(B)** Heatmap of WES analysis demonstrating TMB, INDEL burden, and somatic driver alterations annotated by pre/post-treatment, tumour site, IMDC risk category, and nivolumab response. Composite mutations are annotated with dual colours. Complex mutations in ADR002: *PBRM1* frameshift insertion chr3:52584573:->T and non-frameshift deletion chr3:52584576:TAT>-; *TP53* missense mutation chr17:7572969:A>T and frameshift insertion chr3:7572962:->CT. * (Asterix) denotes two distinct fsINDEL mutations in one tumour sample in ADR013.

## Results

### Patient characteristics and response to nivolumab

15 patients were enrolled from October 2015 to June 2018. Demographic and clinical characteristics are shown in **Table S1A**. 13 (87%) patients had intermediate- or poor-prognostic risk disease (by International Metastatic RCC Database Consortium risk categorisation; IMDC) (**STAR Methods**)^37^. At clinical datalock (December 2018), median follow up was 12.5 months. Six deaths occurred, all due to progressive disease. The median progression-free (PFS) and overall survival (OS) were 4.1 and 12.5 months, respectively. For translational analyses, we defined ‘responders’ as patients who had a partial response (PR) or stable disease (SD) as measured by Response Evaluation Criteria In Solid Tumours criteria (**STAR Methods**) for ≥6 months (five patients). ‘Non-responders’ were classified as patients with progressive disease (PD) within 6 months of enrolment regardless of best-response (ten patients). By these criteria, five patients (33%) had a PR, where one patient (ADR005) had short-lived PR (<6 months, non-responder). Six patients (40%) had SD, where one patient (ADR011) had durable response (>6 months SD, responder) (**Figure S1A and Table S1A**). Two patients underwent a cytoreductive nephrectomy during the course of the study. We observed no associations between age, sex, IMDC risk category, and presence of sarcomatoid/rhabdoid features (n=2) and response to nivolumab (**Table S1A**). Overall, these clinical outcomes data are consistent with a larger phase II (n=110) cohort study of first-line pembrolizumab in patients with mRCC^38^.

### Molecular features do not correlate with nivolumab response

All patients underwent image-guided percutaneous tumour biopsies with additional archived and fresh samples collected via TRACERx Renal and PEACE studies. 15 patients had baseline samples, and 13 post-treatment samples. In total, 115 tumour samples (fresh and archived) were available for translational analyses (**Figure S1A** for consort diagram; **Supplemental Data Table 1** for sample characteristics). 81 fresh tumour samples and matched germline DNA underwent whole-exome sequencing (WES). Subsequently, 21 samples were excluded due to low tumour purity as expected with image-guided biopsies. 59 tumour samples from 13 patients were of sufficient quality for downstream analyses (**STAR Methods**).

Median sequencing depth was 199x (range 130-359x) (**Supplemental Data Table 1**). Neither pre-treatment TMB (median 0.9 mut/mb; range 0.4-11.1), nor fsINDEL load (median 9; range 0-169) associated with response to nivolumab (**Figure S1B**). Reduction of nsSNVs and fsINDELs post-treatment compared to baseline has been referred to as “genomic contraction”^39^, and may reflect immune recognition of neoantigens and subsequent elimination of tumour cells under CPI. To explore the contribution of neoantigens to anti-PD1 response, we asked whether mutations which have undergone genomic contraction post-treatment were enriched for mutations which encoded neoantigens (**STAR Methods**). We found no significant difference in contraction of neoantigen-encoding mutations compared to the remaining non-synonymous mutations (**Figure S1C**).

Molecular features of this cohort were typical of ccRCC^1, 35^, including mutations in VHL (77%, and *VHL* methylation in additional 15%), *PBRM1* (62%), *SETD2* (38%), BAP1 (15%), and *KDM5C* (38%) (**Figure 1B**). Increased sensitivity of multiregion sampling revealed the presence of subclonal driver events in most patients. Instances of composite mutations (two or more nonsynonymous somatic mutations in the same gene and tumour sample^40^) involving *SETD2, KDM5C*, and *TSC2* were detected. There was no association between any mutations, regardless of their clonality, and response to nivolumab, including *PBRM1* (*P*>0.05, Fisher’s exact test). Copy number landscape was also typical of ccRCC with clonal loss of 3p25.3 detected in all tumours and 9p21.3 and/or 14q31.1 loss observed in 12/13 patients, consistent with our previous findings in metastatic ccRCC^36^ (**Figure 1B**). In agreement with other studies, weighted genome instability index (wGII) as a global measure of chromosomal complexity was not predictive of nivolumab response (median 0.29, range 0.04-0.71; P=0.076) (**Figure S1B; STAR Methods**)^19, 20^. No driver SCNAs associated with response while loss of 10q23.31, previously reported to associate with benefit from nivolumab^19^, showed only a trend towards enrichment in responders (P=0.07, Fisher’s exact test). However, the small cohort size was likely statistically underpowered to robustly detect response associations of these mutational events.

Intermetastatic heterogeneity can underpin differential therapy response^41-44^, which we evaluated through post-mortem sampling. ADR015 had stage IV disease at enrolment into ADAPTeR, including surgical bed recurrence, mixed lytic/sclerotic bone, and nodal disease. A tonsillar metastasis was resected pre-treatment. PFS was 8.4 months under nivolumab (best response: PR), patient progressed with multiple brain metastases and died at 27.3 months after trial enrolment (**Figure S2A**). All metastatic deposits were sampled after death and whole exome sequenced. We found evidence of genetic divergence between disease sites which progressed (brain) and responded (nodal metastases) under nivolumab. Mutational profile of a thyroid metastasis incidentally found at autopsy reflected lesions which responded under treatment (**Figures 1B and S2B**). Significantly higher median TMB (10.8 mut/Mb) and fsINDEL load (166) was evident in the progressive brain and resected tonsillar metastases, consistent with a hypermutant genotypic background in these sites, compared with treatment-responsive disease sites (median TMB 1.3mut/Mb; fsINDEL load 8). Accordingly, we observed higher neoantigen load in brain and tonsillar metastases (**Figure S2B**). Excess mutations carried the signature of C>T at Gp<u>C</u>pN trinucleotides (Signature 15) associated with defective DNA mismatch repair^13^. We observed a pathogenic mutation in *MLH1*^45^ with loss of heterozygosity (LOH; biallelic inactivation through canonical 3p loss), in the sites with excess TMB (**STAR Methods**), as well as a beta-2-microglobulin (*B2M*) mutation with LOH (biallelic inactivation through 15q loss) (**Figure S2B; STAR Methods**). Functional *MLH1* mutations result in mismatch repair-deficiency (MMRd) and high immunogenicity driven by accumulation of neoantigens across MMRd tumours^46^. *B2M* encodes a protein subunit integral for major histocompatibility complex class I (MHC-I) endogenous peptide presentation^47^. Taken together, our findings suggest that in this case somatic loss of *MLH1* led to accumulation of an uncharacteristically high number of neoantigens in the context of ccRCC, and subsequent loss of antigen presentation via *B2M* and immune escape, as observed during nivolumab treatment. MMRd in ccRCC has been reported but is rare^48^, and *B2M* loss as a mechanism of immunotherapy resistance^49^ has not to date been described in ccRCC.

### Majority of HERVs detected in ccRCC tumour samples are expressed by immune cells

In light of reports associating intratumoural cytotoxic T-cells^28^ or response to nivolumab^50-52^ with expression of human endogenous retroviruses (HERVs) in ccRCC, we examined previously published HERV signatures in the ADAPTeR cohort. We performed RNAseq on 60 tumour samples, 33 pre-treatment and 27 post-treatment (week-9), representing 14 patients (**Figures S1A; STAR Methods**).

Two of the previous reports of HERV associations^28, 50^ used 66 HERV loci annotated by Mayer et al.^53^, and another study^51^ selected 3,173 HERV loci annotated by Vargiu et al.^54^. We mapped these loci to a custom repeat region annotation which we described previously^55^ (**STAR Methods**). First, we observed discrepancies between our HERV loci annotations with those in Mayer et al. and/or Vargiu et al., including HERV loci that were considered as a single integration in our annotation that appeared fragmented in the Mayer et al. and/or Vargiu et al. annotations, and vice versa (**Supplemental Data Table 2**). Further, we found HERV annotations in both Mayer et al. and Vargiu et al. that were either incomplete or extended beyond the boundaries of integrations to include exons of adjacent genes belonging to separate transcription units (**Figure S3A**). We note that these discrepancies affected HERV integrations previously associated with cytotoxic T-cell presence or ccRCC response to immunotherapy, such as ERV3-2 and ERVK-10^28,51^ (**Figure S3A**). The previously annotated 66 and 3,173 HERVs finally corresponded to 7,989 repeat annotations by our method^55^.

We evaluated previously reported HERV signatures^28, 50, 51, 56^ in our cohort and did not observe a difference between responders and non-responders, nor significant change in their expression levels following nivolumab (**Figure 2A**). Further, we found that the HERVs with the highest expression and, therefore, strongest contribution to previously described signatures^28, 50, 51, 56^, such as ERV3-2 and ERVK-10, were not specific to ccRCC, but were highly expressed in purified immune cell subsets (**Figure S3B; STAR Methods**). Therefore, it seems likely that their reported association with response to CPI is underpinned by immune infiltration (which in itself is linked to response^20, 21^) and high HERV expression in immune cells.

**Figure 2.**
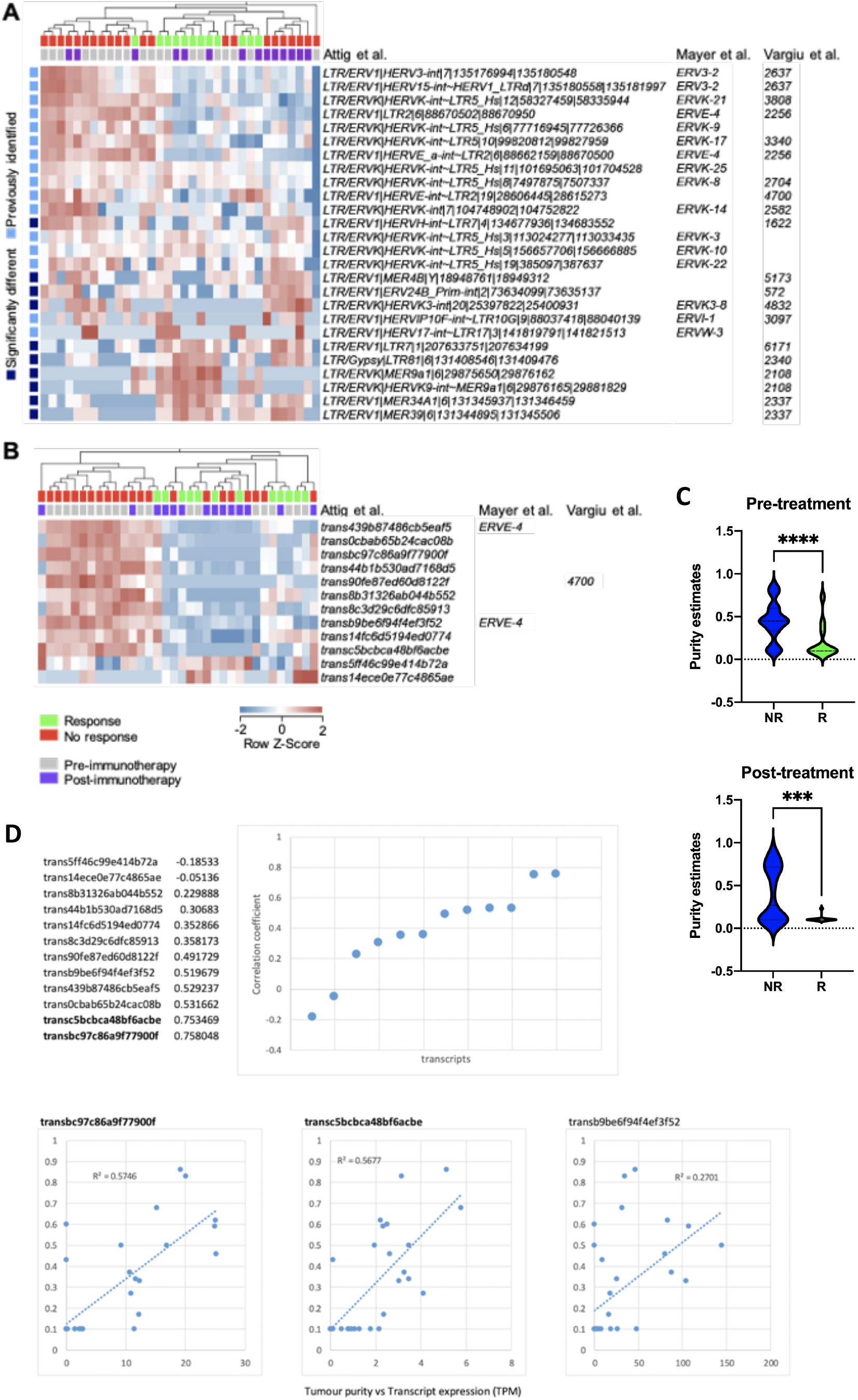
Expression of human endogenous viruses (HERVs) and LTR-overlapping transcripts in ccRCC according to tumour purity. **(A)** Hierarchical clustering patient samples according to the relative expression of HERVs previously associated with cytotoxic T-cell presence, response to immunotherapy or the provision of antigenic epitopes. **(B)** Hierarchical clustering patient samples according to the 12 LTR-overlapping transcripts that were differentially expressed (≥2-fold change, q≤0.05) between responders and non-responders or affected by nivolumab. **(C)** Comparisons of tumour purity between non-responders and responders. Per-sample values are represented. Median values are shown. \*\*\*\**P*<0.0001, \*\*\**P*=0.001; Mann-Whitney U test. **(D)** Distribution plot of significant Spearman’s rank-order correlation between tumour purity and transcript per million (TPM) expression of the 12 HERVs differentially expressed between responders and non-responders. *NR - non-responders; R - responders*

Next, to examine a possible correlation with HERVs that were ccRCC-specific, we measured expression of 570 transcripts previously identified through *de novo* transcriptome assembly to overlap with LTR elements and with high specificity to ccRCC^57^ (**STAR Methods**). Most ccRCC-specific LTR elements were expressed (≥0.5 TPM) in the majority of samples in this study and 12 ccRCC-specific LTR elements, from nine distinct loci, were differentially expressed (≥2-fold change, q≤0.05) between response groups or their expression levels were significantly altered following nivolumab (**Figure 2B**). These transcripts included members of the HERV-E group (ERVE-4^53^ and HERV4700^51^) that were previously associated with immunotherapy response in ccRCC^51-53^. However, we found that these transcripts were expressed predominantly in pre-treatment non-responders (**Figure 2B**). Tumour purity in pre-treatment samples was significantly higher in non-responders compared to responders (reflecting lower levels of immune infiltration) (**Figure 2C**). Consequently, the level of HERV expression correlated with tumour purity (**Figure 2D**), explaining higher expression in non-responders. Post-treatment, we observed that the expression of ccRCC-specific LTR transcripts in non-responders normalised relative to responders (**Figure 2D**). A possible explanation is that nivolumab-induced immune infiltration lowering tumour purity, which in turns lowered the abundance of tumour-specific LTR transcripts in the biopsies.

In summary, although we did observe correlations between expression of certain HERVs and response to nivolumab in our cohort, as described in earlier studies^28, 50, 51, 56^, we further found that these correlations were indirect. Firstly, certain HERVs, such as ERV3-2 and ERVK-10, were associated with immunotherapy response in ccRCC owing to their high expression in immune cells, rather than tumour cells. Secondly, abundance of ccRCC-specific HERV transcripts, such as ERVE-4 and HERV4700, reflected tumour purity and correlated with response to immunotherapy indirectly as the least infiltrated tumours responded the least to therapy.

### Nivolumab induces T-cell activation and upregulation of TCR signalling in responders

Next, we evaluated the composition of the immune infiltrate in the bulk transcriptome of tumour samples pre- and post-nivolumab. We performed differential gene expression, gene set enrichment analysis (GSEA), and immune subset deconvolution^58^ (**STAR Methods**). Immune infiltration and cytotoxicity signatures were observed in all tumours and all time points, but their expression was significantly higher in responders compared to non-responders (*P*=0.031 and *P*=0.017, respectively), and higher post-treatment compared to pre-treatment in both groups (**Figures 3A-3D**). Treatment-induced enrichment of “immune-activation” and the “TCR signalling” pathways was evident in responders but not in non-responders (**Figures 3E and 3F**). Specifically, using the Danaher signature^58^, the T-cell score expression was significantly higher in responders at pre- and post-treatment timepoints (*P*=0.019 and P=0.038 respectively) (**Figure S4**). We also observe a trend towards higher B-cell expression scores in responders pre-treatment, but not post-treatment (Figure S4).

**Figure 3.**
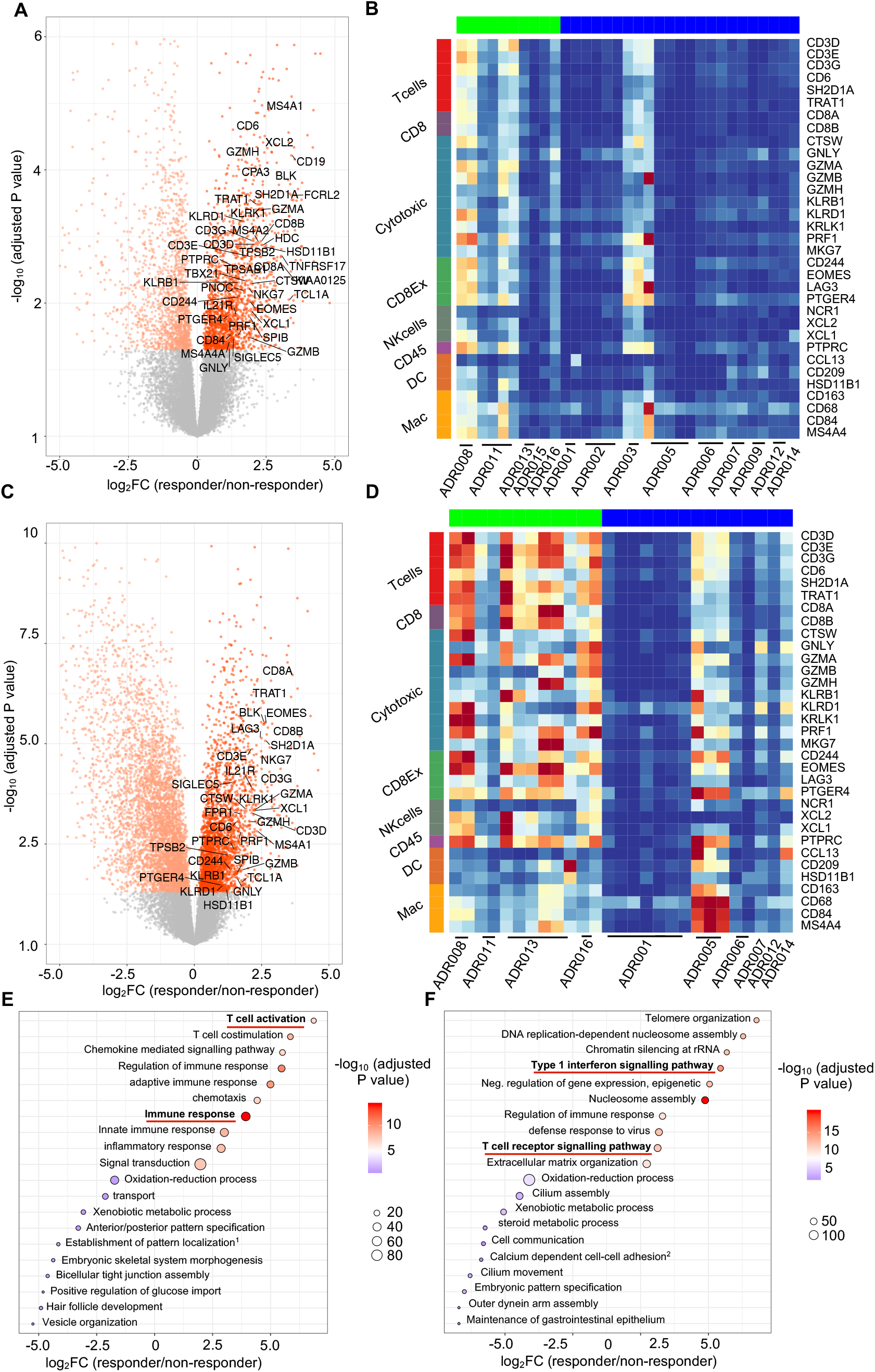
GSEA and immune deconvolution by RNAseq shows higher levels of immune infiltration and activation in responders compared to non-responders under nivolumab. **(A)** Transcripts differentially regulated Pre-treatment between responders and non-responders (n=33 samples, 14 patients, negative binomial Wald test, Benjamini–Hochberg corrected *P* values). 3,382 transcripts were differentially regulated (FDR<0.05), the ones that overlap with the Danaher immune score gene list are labelled. **(B)** Heatmap showing the relative expression (z scores) of genes from 8 Danaher immune modules in Pre-treatment samples. **(C)** Transcripts differentially regulated post-treatment between responders and non-responders (n=27 samples, 10 patients, negative binomial Wald test, Benjamini–Hochberg corrected *P* values). 7,975 transcripts were differentially regulated (FDR<0.05), the ones that overlap with the Danaher immune score gene list are labelled. **(D)** Heatmap showing the relative expression (z scores) of genes from 8 Danaher immune modules in post-treatment samples. **(E)** GOBP pathway analysis of genes preferentially upregulated and downregulated pre-treatment in responders, Overlap (n), number of significant genes from a pathway (hypergeometric test). **(F)** GOBP pathway analysis of genes preferentially upregulated and downregulated post-treatment in responders, Overlap (n), number of significant genes from a pathway (hypergeometric test).

Finally, we evaluated the association between previously published associations between gene expression signatures and CPI response in metastatic ccRCC (**STAR Methods**). IMmotion150 study T_eff_ ^high^ signature^20^, but not T_eff_ ^high^/Myeloid^low^ signature was enriched in responders compared to non-responders (P=0.042 and P=0.038 at pre- and post-treatment timepoints, respectively) (Figure S4). The 26-gene Javelin101 signature^21^ was also enriched in responders compared to non-responders (P=0.028 and P=0.038 at pre- and post-treatment timepoints, respectively). Individual marker genes for T-cell infiltration (CD3E, CD8A), activation (GZMB), and TCF7 expression (reported as predictive of CPI response^52^), were consistently higher in responders compared to non-responders, particularly post-treatment (Figure S4). Overall, previously published signatures of response validate in our cohort in spite of inherent differences in treatment regimens, and tissue type used for transcriptome profiling.

### CD8^+^ T-cells upregulate GZMB following nivolumab in responders

Following antigen stimulation CD8^+^ T-cells undergo cytotoxic differentiation in order to mediate tumour cell killing. To investigate the phenotype of the T-cells in the tumours under study, we performed multiplex immunohistochemistry (mIHC) and immunofluorescence (mIF) on 61 formalin-fixed paraffin-embedded tumour samples (41 pre-treatment; 20 post-treatment) from 14 patients (**Figure S1A**). We applied bespoke antibody panels (3 markers for mIHC, 6 markers for mIF) to quantify and characterise infiltrating immune cells (**STAR Methods**).

We observed no difference in T-cell number (CD8^+^, CD4^+^, CD8^+^CD4^+^, or Tregs), or CD8^+^/Treg and CD4^+^effector/Treg between response groups, either pre- or post-treatment (Figures 4A and S5A-C). Total PD1 expression did not differ in the two groups (**Figures 4B**). Overall quantified GZMB expression (*P*=0.024) and GZMB expression on CD8^+^ T-cells (*P*=0.047) were significantly higher in responders compared to non-responders post-treatment, but not pre-treatment (**Figure 4B**). Where paired pre- and post-treatment tumour samples were available, we observed a trend for upregulation of GZMB expression on CD8^+^ T-cells in responders but not in non-responders, following nivolumab (**Figure S5D**). CD163^+^ myeloid cell level alone or as a ratio to T-cells (CD3^+^/CD163^+^ and CD8^+^/CD163^+^) did not associate with response (**Figures 4A and S5C**). Compared to non-responders we observed significantly more B-cells in responders pre-but not post-treatment (*P*=0.02 and 0.96, respectively) (**Figures 4A**). There were no differences in the number of plasma cells between response groups (*P*=0.23 and 0.54 pre- and post-treatment, respectively) (**Figures 4A and S5A**).

**Figure 4.**
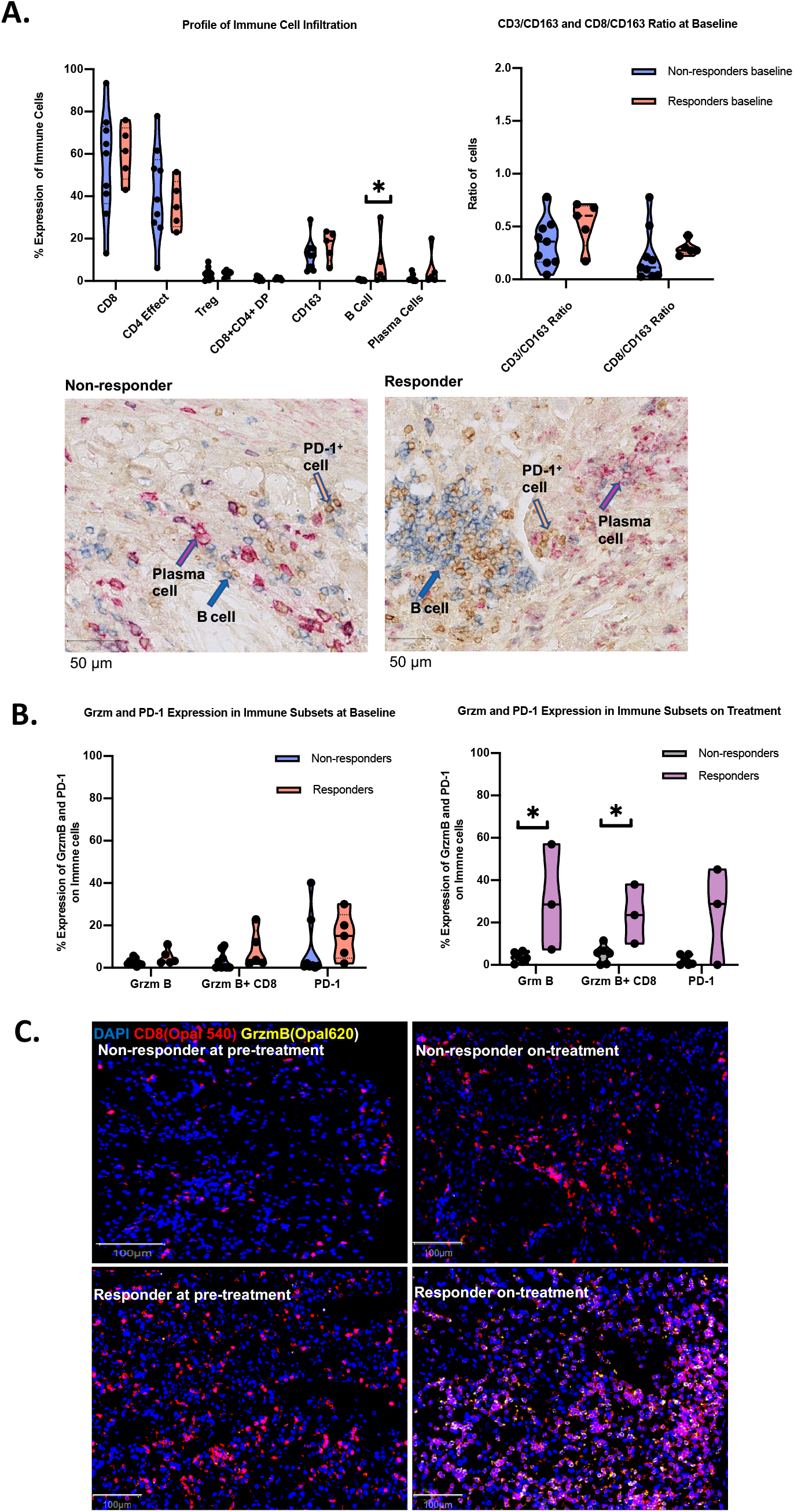
Quantification and immunophenotyping of pre- and post-treatment infiltrating immune cells by IHC and mIF. **(A)** Comparison of T-cell subset (out of total T-cells), CD163^+^ myeloid cells, B-cell and plasma cell infiltration in treatment naïve samples in responders (n=5) and non-responders (n=9) is shown on the left. On the right is the ratio between CD3 (total T-cells) and CD163 myeloids cells and CD8 and CD163 cells at baseline. B-cell and plasma cell scoring was done by using immunohistochemistry. Other markers were scored by using IF. IHC images of representative responder and non-responder patients pre-treatment showing B-cell (blue), PD-1^+^ cells (yellow) and plasma cells (magenta) infiltration. **(B)** Level of overall GZMB, GZMB^+^CD8^+^, and overall PD-1 expression in responders and non-responders in treatment naïve and on treatment samples is shown. PD-1 staining was performed with immunohistochemistry. All other markers were stained with IF. **(C)** mIF images showing GZMB^+^CD8^+^ cells in a representative responder and non-responder patient at baseline and on Nivolumab treatment. Median values were used for each patient and a two-sided Mann-Whitney U statistical test was used for the analysis.

We note a certain number of discrepancies in our observations made from bulk RNAseq and mIHC/IF data. For example, increased B-cells and higher GZMB expression in responders evident by mIHC/mIF were non-significant trends by RNAseq (**Figure S4**), and the opposite trend was observed for CD4^+^/8^+^ T-cell numbers and PD-1 expression. These findings likely reflect the known weak correlation between protein and mRNA levels for many genes, as well as sensitivity limitations of immune deconvolution and classification by bulk RNAseq^60^ as compared to the single-cell resolution afforded by histology-based methods.

### Maintenance of TCR clonal expansion and clustering support ongoing antigen-driven stimulation of T-cells in responders

Next, we sequenced the β-chain TCR repertoires from 64 tumour and 29 peripheral blood mononuclear cell (PBMCs) samples from 14 patients pre- and post-treatment (**Figure S1A; STAR Methods**). To mitigate against effects of intratumour TCR heterogeneity^61-63^ (**Figures S6A and S6B**), we pooled TCR sequences from multiple tumour regions for each patient at different timepoints.

Cohort-wide median number of unique β-chain transcripts in tumour and blood samples was 3,644 and 21,370, respectively. For each pooled sample, we quantified TCR diversity through a TCR repertoire clonality score (low scores correlate with more diverse repertoire and high scores with dominant clones) (**STAR Methods**). TCR clonality was overall higher in tumour samples compared to PBMCs (**Figure 5A**), potentially reflecting intratumoural expansion of specific TCR clones. T-cell clonal contraction and expansion was not significantly different between tumour and PBMCs, irrespective of response (**Figures 5B and S6C**,**E**). Contraction and expansion of CDR3s present in pre- and post-treatment samples was not associated with response. This was true for tumour and PBMC samples (**Figures 5C and S6D**). To examine the maintenance of expanded clonotypes, we computed a cosine score which reflects degree of TCR repertoire similarity comparing pre- and post-treatment timepoints (**STAR Methods**). Compared to non-responders, intratumoural, but not PBMC TCR repertoire similarity was significantly greater in responders (*P*=0.024, **Figures 5D and S6F**). In responders, intratumoural clones expanded pre-treatment were more likely to be maintained compared to non-responders, where they were frequently replaced by new clones (*P*=0.024, **Figures 5E and S7A**).

**Figure 5.**
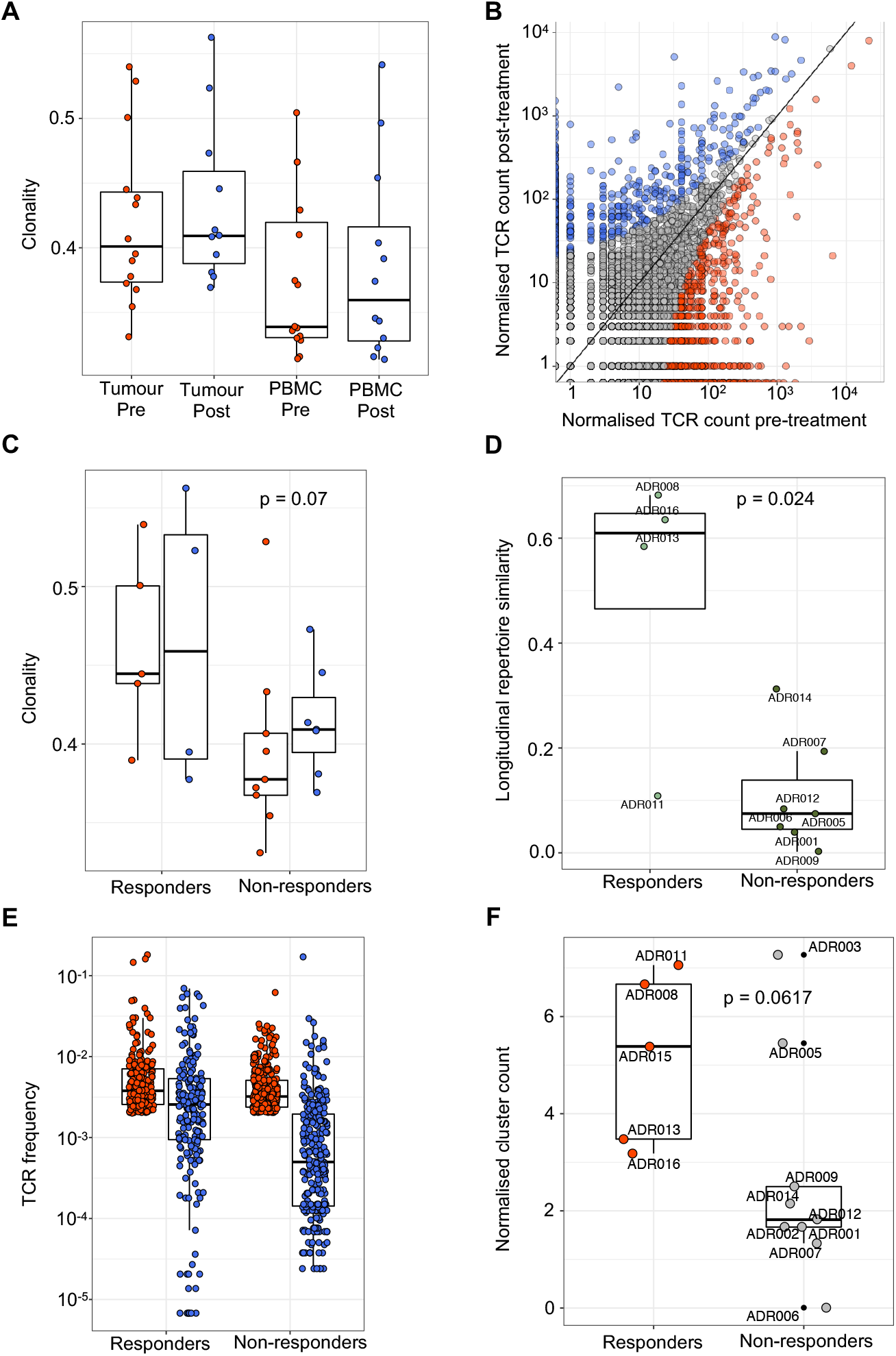
TCR sequencing demonstrates maintained clonal expansion through persistent antigenic stimulation associate with nivolumab response. **(A)** The intratumoural and peripheral TCR repertoire clonality score is shown for each patient at each timepoint. **(B)** Correlated clone sizes in tumour samples. Scatter plots of tumour clone size after treatment and before treatment are shown for all patients. Clones are colored by expansion/contraction status (**STAR Methods**). **(C)** The intratumoural TCR repertoire clonality score pre-treatment and on-treatment is shown for each patient. Patients are split between responders and non-responders. Mixed-effect model p-value shown. **(D)** The intratumoural cosine score between pre-treatment and on-treatment is shown for each patient (n=12). Patients are split between responders and non-responders. Responding patients exhibit greater cosine score, with the two-sided Mann–Whitney test *P* value shown. **(E)** The frequency distribution of the intratumoural expanded TCRs pre-treatment (red circles; *n* = 469 individual TCRs combined from 12 patients) and post-treatment (blue circles). Only TCRs that were detected post-treatment were included. **(F)** The clustering algorithm was run on all patients, and the pre-treatment normalised number of clusters for the networks containing expanded sequences is shown. Two-sided Mann–Whitney test *P* value shown; n=14 patients.

Antigen specific T-cell responses are often associated with the presence of clusters of TCRs with similar CDR3 peptide binding sequences^64, 65^. To investigate if maintained expansion of TCR clonotypes is driven by a shared and persistent antigen, we performed clonotype clustering analysis (**STAR Methods**). In responders, expanded TCR clones exhibited a trend towards increased clustering of similar CDR3 sequences (or ‘cluster structure’) compared to non-responders both pre- and post-treatment (*P*=0.06 and 0.07, respectively) (**Figure 5F and S7F**). Expanded-maintained TCRs displayed significantly more cluster structure than expanded-replaced TCRs (*P*=0.008, **Figure S7C-E**). Together, these data suggest that in responders TCR clonotypes expanded due to a shared antigen and were maintained presumably due to persistent antigen stimulation.

Finally, we performed TCRseq in a patient who underwent post-mortem sampling (ADR005), allowing greater resolution of spatial and temporal dynamics of TCR clonotypes and insight into surveillance of metastatic lesions at the time of death. While primary tumour and lung metastases in this case maintained a partial response to nivolumab until death, new brain, bone, and thoraco-nodal metastases emerged on nivolumab, presenting sites of immune-escape (**Figure S2C**). Five TCR clones that were expanded pre-nivolumab were maintained post-treatment at week-9. Three of these five clones remained expanded in non-progressive disease sites (lung metastasis and primary tumour), but none were detected in the sites of progression (brain, bone and thoraco-nodal metastases) (**Figures S2D and S2F**).

### Responder CD8^+^ T-cells express TCF7

TCR stimulation drives T-cell differentiation states which impact effector function^66-68^ and response to CPI^69^. We derived single-cell suspensions of tumour-infiltrating lymphocytes from six spatially distinct regions of the nephrectomy specimen and analysed CD8^+^ T-cell differentiation states via high dimensional flow cytometry (**STAR Methods**). Due to large amounts of fresh tissue required, this was only feasible in the patients who underwent cytoreductive nephrectomies (ADR013, responder; ADR001, non-responder).

TCF7 is associated with progenitor-like phenotype and preserved effector function of dysfunctional T-cells during prolonged antigen stimulation^67, 70, 71^. A higher frequency of CD8^+^ T-cells expressed TCF7 in the responder ADR013 (27.5%) compared to non-responder ADR001 (4.64%). TOX expression is linked with dysfunction, but also appears critical for epigenetic programming of progenitor-like CD8^+^ T-cell towards effector function^72-75^. More TCF7^+^TOX^+^CD8^+^ T-cells were detected in ADR013 compared to ADR001 (4.56% vs. 0.96%, respectively) (Figures 6A-C). CD39 associates with antigen stimulation and tumour reactivity^76, 77^. CD39 expression was higher in ADR013 (35.7%) compared to ADR001 (2.29%) (**Figures 6A-C**). Expressed markers of dysfunction^78, 79^ in ADR013 and ADR001 were: PD-1 (29.2% and 2.25%), TIM3 (13.1% and 1.35%), and CD38 (33.9% and 19.7%), respectively (Figures 6B-C). The markers differentiating ADR013 from ADR001 were reflected in the bulk RNAseq cohort-level data, which showed a trend towards higher expression of TCF7, CD39, and TOX in responders compared to non-responders (P=0.11, 0.35, and 0.07, respectively) (**Figure S4**).

**Figure 6.**
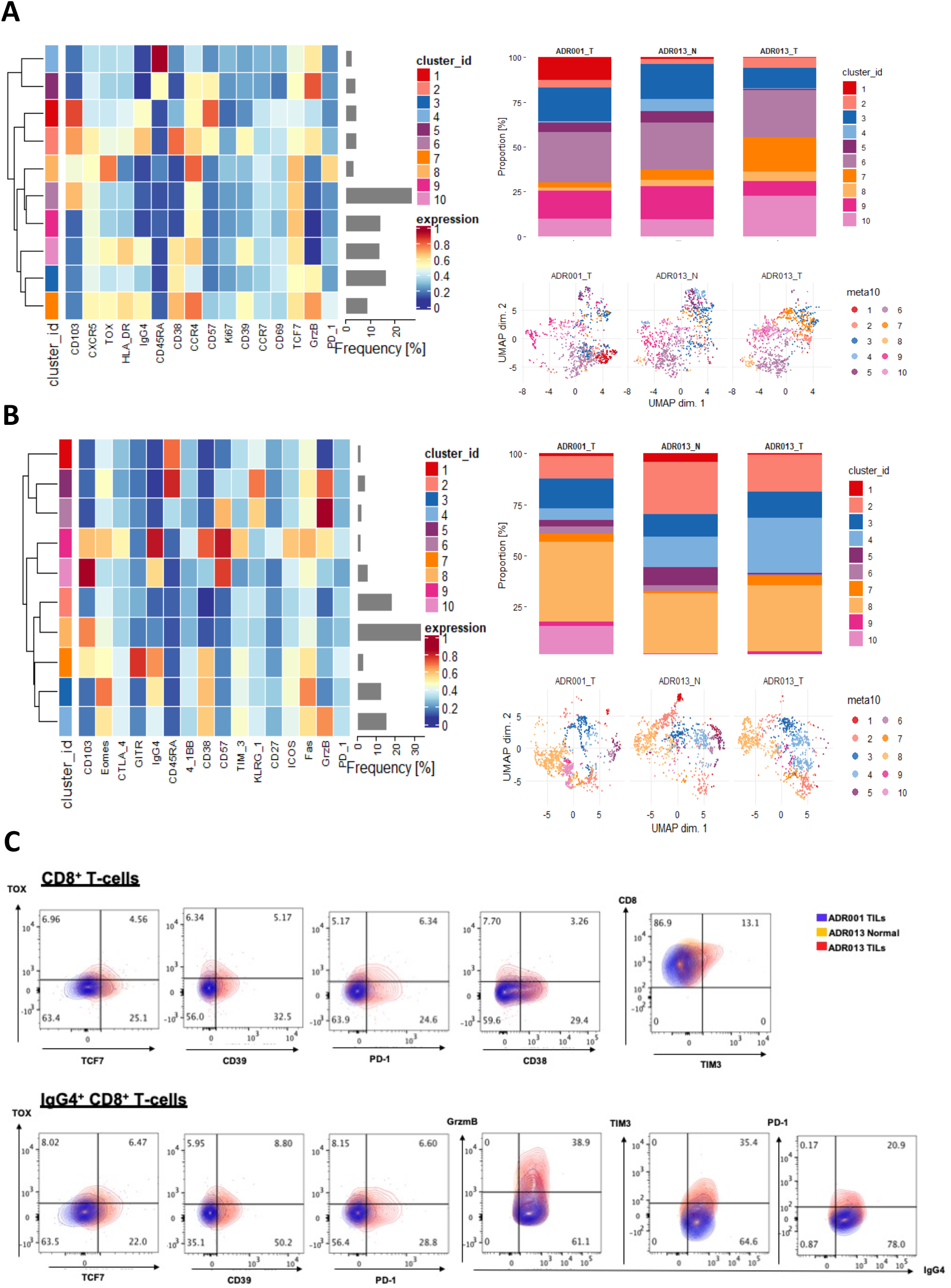
Flow cytometry-based analysis of ADR013 (responder) and ADR001 (non-responder) evaluating post-treatment total and nivolumab-bound CD8^+^ T-cells. **(A)** Expression of T cell differentiation focused markers on CD8^+^ tumour infiltrating lymphocytes in a representative patient who had ≤6 months response to Nivolumab treatment (ADR001_T tumour tissue) and a representative patient who had had ≥6 months response to Nivolumab treatment (ADR013_T tumour tissue and ADR013_N tumour adjacent normal kidney tissue) is shown in the heatmap. Relative expression level of each heatmap cluster for each sample is shown in the percentage bar graph and UMAP. 920 CD8^+^ cells were used per sample for the analysis. **(B)** Expression of T cell checkpoint focused markers on CD8^+^ tumour infiltrating lymphocytes in a representative responder and a non-responder patient is shown in the heatmap. Relative expression level of each heatmap cluster in each sample is shown in the percentage bar graph and UMAP. 990 CD8^+^ cells were used per sample. **(C)** FACS plots show the co-expression of markers on CD8^+^ and IgG4^+^CD8 cells in ADR001_T tumour, ADR013_T tumour tissue and ADR013_N tumour-adjacent normal kidney tissue.

### Expanded CD8^+^ T-cells are drug-binding and cytotoxic during nivolumab induced responses

Next, we asked if characterising intratumoural, nivolumab-bound cells and comparing responder and non-responder populations would provide further resolution on CD8^+^ T-cells which exhibit features of antigen engagement. IgG4 has previously been shown as a robust surrogate marker for PD-1 receptor occupancy by anti-PD1 antibodies^80, 81^. We established the technical feasibility of this method in a competition assay where IgG4 identified T-cells bound to pembrolizumab (anti-PD1 antibody), consistent with prior reports (**Supplemental Data Figure 1; STAR Methods**).

We performed flow cytometry analysis on IgG4^+^CD8^+^ T-cells from ADR0013 (responder) and ADR001 (non-responder). Expression of the following markers were all higher in ADR013 compared to ADR001: GZMB (38.9% vs 8.75%), TCF7 (19.5% and 2.17%), CD39 (54.6% vs 3.25%), TOX (14.5% vs 4.10%), and TIM3 (35.4% vs 3.52%), respectively (**Figure 6C**). This suggested drug-bound CD8^+^ T-cells in the responder were cytotoxic and progenitor-like, despite upregulating markers of dysfunction. We detected unbound PD-1 on IgG^+^CD8^+^ T-cells despite expected receptor occupancy by nivolumab, higher in ADR013 (20.9%) than ADR001 (0.78%) (**Figure 6C**). Detection of unbound PD-1 on IgG4^+^CD8^+^ T-cells possibly indicates further PD-1 upregulation after drug-binding and TCR stimulation, as a marker of activation^82, 83^, rather than incomplete receptor-drug occupancy in this case.

Next, we performed paired single-cell RNA and TCRseq (scRNA/TCRseq), on IgG4^+^ and IgG4^-^ CD3^+^ T-cells (**STAR Methods**). The scRNA datasets were annotated with their corresponding VDJ information, merged, and followed by UMAP projection. Cells were classed as CD8 (CD8^+^CD4-FOXP3-), CD4 (CD8-CD4^+^FOXP3-) and Treg (CD8-FOXP3^+^) (**Figure S9A; STAR Methods**). We observed similar levels of CD8^+^ T-cells, but lower proportions of Tregs in ADR013 (responder) compared with ADR001 (**Figure S8B**). Differential gene enrichment and gene set enrichment analyses of IgG4^+^CD8^+^ T-cells showed upregulated pro-inflammatory cyto/chemokine genes and T-cell activation pathways in both patients, indicating drug-binding CD8^+^ cells had similar transcriptional characteristics irrespective of clinical response (**Figures 7A and S9C**). TCRseq revealed hyperexpanded CD8^+^ T-cells (defined as 200-1000 clones with the same CDR3 sequence) in ADR013 but not ADR001, where expansions were restricted to <200 clones (**Figures 7B and 7C**). We observed that the more pronounced the clonal expansion, the higher the proportion of IgG4^+^ compared to IgG4^-^ CD8+ T-cells, suggesting drug-binding led to clonal expansion (**Figure 7D**). Expanded IgG4^+^CD8^+^ clones were characterised by cytotoxic activation in ADR013, including higher expression of GZMK than ADR001 (**Figures 7E and S9D**). Analysis with a portfolio of publicly available gene signatures for T-cell states demonstrated expression of signatures associated with T-cell activation / dysfunction in drug-bound cells from both patients, higher in ADR013 than AD001, consistent with increased TCR stimulation in the responder and in drug-bound cells (Figure S9E; **STAR Methods**). scRNAseq data also confirmed flow cytometry findings, with higher expression of GZMB, TCF7, TIM3, and CD39 expression on IgG4^+^CD8^+^ T-cells in ADR013 compared to ADR001 (Supplemental Data Figure 2).

**Figure 7.**
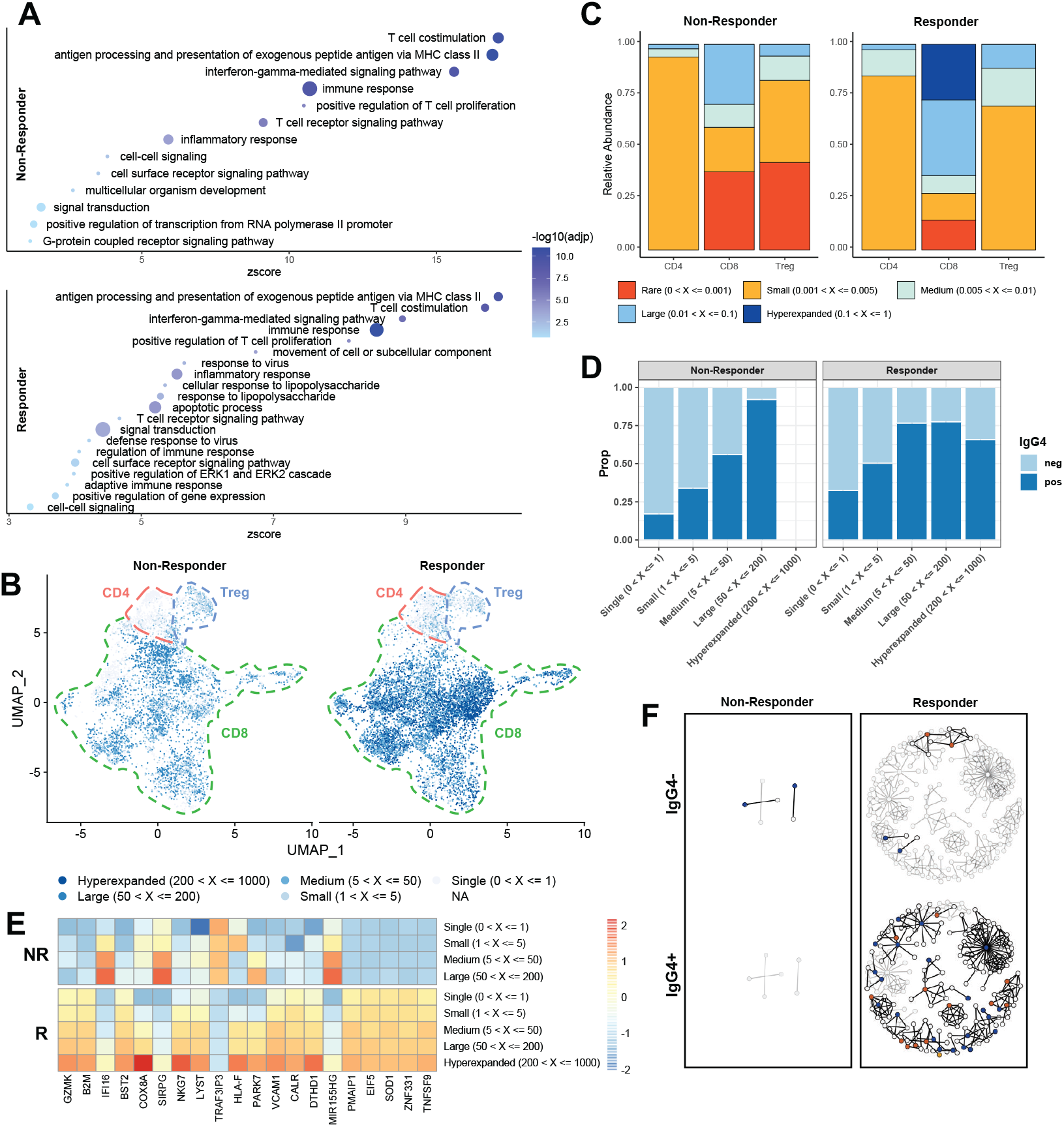
Nivolumab binding correlates with upregulation of T-cell activation genes and clones expanded through persistent antigenic stimulation. **(A)** GOBP pathway analysis of genes preferentially upregulated in drug bound CD8 cells in ADR001 (non-responder) and ADR013 (responder), circle size indicative of number of genes overlapping with GOBP term. **(B)** UMAP of scRNA Seq data from non-responder and responder coloured by frequency of clone. **(C)** Clonal proportion plot of CD8, CD4 effector and Treg compartments in non-responder and responder. **(D)** Proportion of cells in each expansion class which are Nivolumab bound or unbound. **(E)** Heatmaps showing top genes which positively correlated (Pearson’s correlation, CD8^+^ cells only) with TCR expansion in the responder. **(F)** Representative network diagrams of post-treatment intratumoural CDR3 β-chain sequences for ADR001 and ADR013. Clustering was performed within the bulk TCR-seq data around expanded intratumoural TCRs, subdivided between clones that were expanded in the post-treatment repertoire exclusively (blue circles) and clones that were also expanded Pre-treatment (orange circles). The network shows clusters for which at least one CDR3 was also detected in the scTCR repertoire. IgG4 negative clones that were detected in the scTCR repertoire but not expanded in the bulk TCR repertoire and are represented (yellow circle). The network was then split between clones that were mapping to a majority of IgG4 negative cells (top panel) or a majority of IgG4 positive cells (bottom panel) in the single-cell data. Clustering network derived from bulk post-treatment tissue (grey circles) as also shown in **Figure S7G**.

Next, combining bulk-TCRseq and scTCRseq datasets, we asked if post-treatment expanded clones in each patient 1) displayed cluster structure (shared antigen recognition); 2) if clustered clones were drug-bound; and 3) if clustered, drug-bound clones were ‘novel’ or from a ‘pre-existing’ population. First, we constructed cluster networks for ADR013 and ADR001 according to CDR3 amino acid triplet sharing (**STARMethods**). Then, we defined each TCR clone within the networks by drug-binding status (IgG4^+^ or IgG4-). Finally, we used pre/post-treatment bulk-TCRseq data for ADR013 and ADR001 to derive ‘novel’ or ‘pre-existing’ labels for each clone which was captured by scTCRseq. The results were scTCR cluster networks, with expanded clones annotated for drug-binding and ‘novel’ versus ‘pre-existing’ status, for each patient (**Figure 7F**). In ADR013 (responder), expanded clones were clustered and mostly (89%) drug-bound. These clustered TCR clusters contained both novel and pre-existing TCRs (**Figure 7F**). By contrast, the salient finding in ADR001 (non-responder) was an overall paucity of expanded or clustered TCRs, either novel or pre-existing, consistent with the post-treatment bulk-level data (**Figure 7F**). This limited any inference on the relationship between clustering and drug-binding at single-cell level in this non-responder patient.

Overall, while the scRNA/TCRseq data were derived from only two patients, not only were the findings in agreement with the bulk, cohort-level data, showing maintenance and reinvigoration of pre-existing, progenitor-activated CD8^+^ T-cells underpinned nivolumab response; but these data provided direct evidence that intratumoural T-cells in a responding patient were expanded, PD1 expressing and nivolumab-binding, and had a more activated phenotype. These features were not observed in T-cells from a non-responder (**Figure 8**).

**Figure 8.**
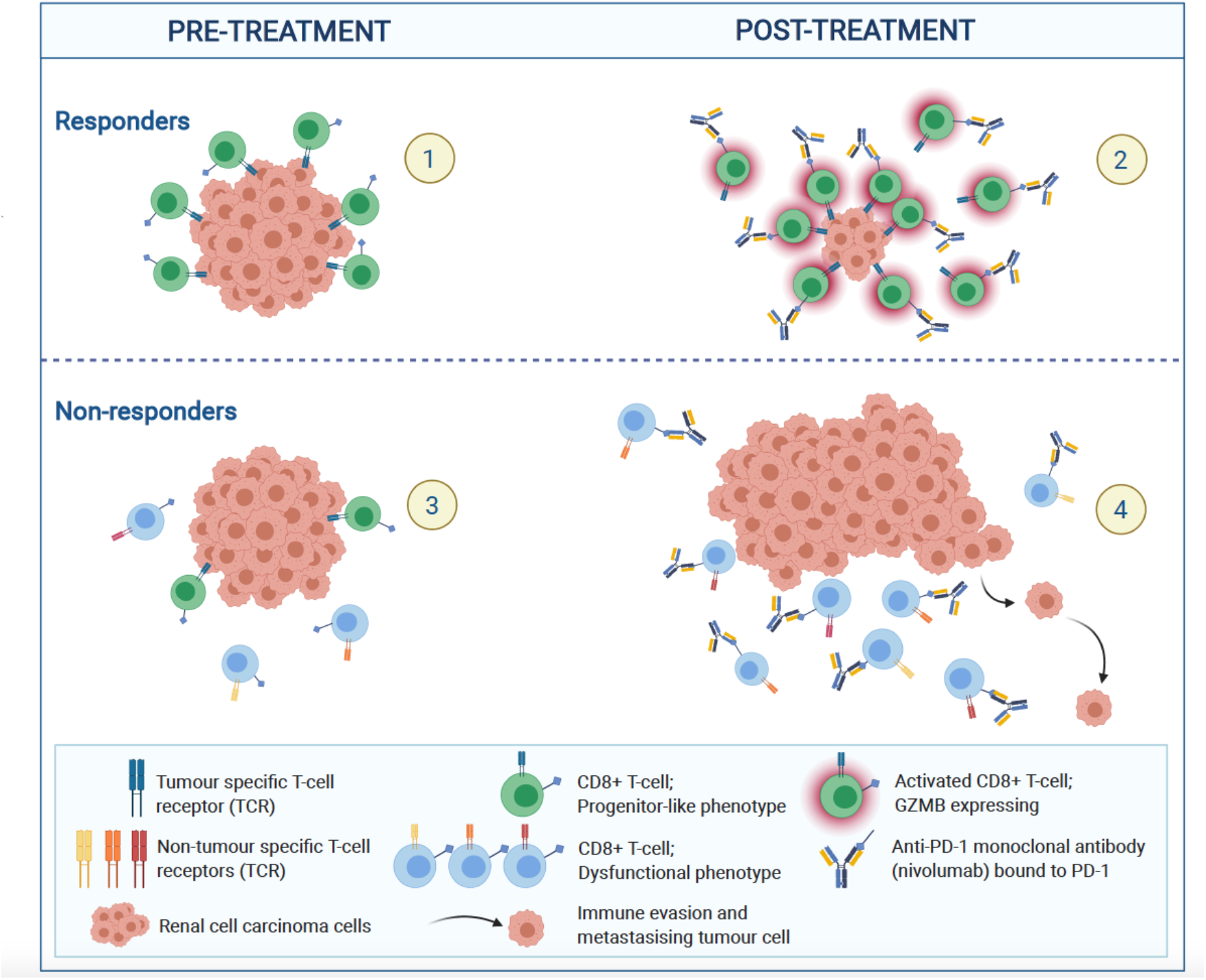
Longitudinal profiling by bulk and single-cell RNA/TCRseq reveal dynamic immune correlates of response and resistance to nivolumab. **(1)** Clonally expanded CD8^+^ T-cells pre-treatment in ADR013 (responder). TCR clonotypes are highly similar. **(2)** Maintenance of pre-existing clonally expanded and expansion of novel CD8^+^ T-cells under nivolumab. Drug-binding activates tumour-specific CD8^+^ T-cells during therapy response. **(3**) Limited clonal expansion of CD8^+^ T-cells pre-treatment in ADR001 (non-responder). TCR repertoire clonality is limited. **(4)** Clonal replacement of expanded CD8^+^ T-cells under nivolumab. Drug-binding occurs on non-tumour specific CD8^+^ T-cells and tumour progression ensues.

## Discussion

We present a multi-omic analysis of advanced stage ccRCC through treatment which sheds light on the mediators of anti-PD1 response and resistance, and in particular the nature of the CD8^+^ T-cells that contribute to anti-tumour immunity.

Our WES data showed that no single mutation, SCNA, nor TMB and INDEL load associated with response in accordance with prior studies^19-21, 27^, although the question about the contribution of mutations or SCNA events to anti-tumour immunity in ccRCC remains incompletely understood. A notable exception was a case with excessively high TMB mediated by MMRd where immune editing was evident with subsequent immune escape via loss of *B2M*. Decreased MHC-I expression associates with reduced PFS with avelumab plus axitinib in ccRCC^21^, but the frequency and impact of loss of antigen presentation remains unclear. With respect to HERV expression signatures, we show that HERVs, such as ERV3-2 and ERVK-10, most frequently associated with T-cell infiltration in bulk tumour biopsies^28, 50, 51, 56^ are highly expressed in immune cells, which offers a more parsimonious explanation for previously described associations. However, we confirmed ERVE-4 and HERV4700 are ccRCC-specific consistent with studies demonstrating direct T-cell reactivity to these specific HERVs^56^. While they did not associate with nivolumab response in this cohort, we note that T-cell responses targeting these HERVs are HLA-A*02 restricted^51, 84^ and, consequently, a positive correlation with the outcome of immunotherapy would only be expected in patients with this HLA allele.

While the source of antigen(s) stimulus in ccRCC remains elusive, their existence is supported by our findings of a population of pre-existing, expanded CD8^+^ T-cells in responders. Moreover, our data show the quality of T-cells were comparable between patients at baseline, but the ability of expanded CD8^+^ T-cells to be maintained underscores response to nivolumab in ccRCC. We show that on-treatment change in GZMB expression is a dynamic biomarker of nivolumab in ccRCC, which has also demonstrated predictive utility for neoadjuvant avelumab in bladder cancer^85^. Increase in TCF7^+^CD8^+^ T-cells and B-cells also correlated with response in our cohort. We note a prior report has shown TCF7^+^CD8^+^ T-cell can be activated *in vitro*, and could maintain a progenitor-like state when located within antigen presentation niches in ccRCC^85^. Higher CD8^+^ T-cell density at tumour invasive margin has been reported to associate with longer PFS with avelumab plus axitinib in ccRCC^21^. As such, further work to characterise the interaction between co-located B- and T-cells, especially at tumour margins will be critical.

There are limitations to our study. First, the small number of patients may limit data generalisability. However, our scope for discovery was afforded by a broadened sampling frame (multiregion and multi-metastatic site biopsies) and longitudinally tracking of molecular and tumour immune microenvironment (TIME) changes under therapy. Only two patient samples underwent multiparameter flow cytometry and scRNA/TCRseq analyses in our study. While this facilitated high-resolution cellular characterisation, spatial relationship with other immune cells was not evaluable. Looking forward, spatial transcriptomic profiling techniques with single-cell sensitivity^87, 88^ will be valuable in studying TIME evolution in ccRCC.

In conclusion, in this prospective study we reveal features of anti-PD1 response and resistance in ccRCC. We identified antigen-specific T-cells with cytotoxic features in ccRCC, which hold promise for development of adoptive cellular therapy for this cancer^89^. While the treatment landscape has evolved to include combination therapies^11^, this dissection of immune changes under nivolumab provides the foundation for understanding response to combination therapies. Finally, our multi-omic analysis framework provides a template and highlights challenges for future immuno-oncology biomarker studies in ccRCC.

## Supporting information

Suppl Item Table 1

Suppl Item Table 2

Suppl Item Table 3

## Data Availability

Further information and requests for resources, data and reagents should be directed to and will be fulfilled by the Lead Contact, Samra Turajlic.

## Acknowledgements

We thank the ADAPTeR trial team and the Skin and Renal Unit Research Team at The Royal Marsden NHS Foundation Trust, including Lyra Del Rosario, Karla Lingard and Mary Mangwende, as well as Kim Edmonds, Sarah Sarker, Charlotte Lewis, Fiona Williams, Hamid Ahmod, Eleanor Carlyle, Tara Foley, Dilruba Kabir, Justine Korteweg, Aida Murra, Nahid Shaikh, Kema Peat, Sarah Vaughan and Lucy Holt. We acknowledge the valuable support of the PEACE consortium. We also thank Lavinia Spain, Irene Lobon, Daqi Deng, Katja De Paepe, Andy Georgiou, Carmella Beastall, Nagina Mangal, Katey Enfield, and Dhruva Biswas for their input. Most importantly, we thank the patients and their families.

The ADAPTeR study (CA209-129) is sponsored by The Royal Marsden NHS Foundation Trust, and partly funded by National Institute for Health Research (NIHR) Biomedical Research Centre (BRC) at the Royal Marsden Hospital and Institute of Cancer Research (ICR) (A80), and Cancer Research UK (CRUK) (17767). Bristol-Myers Squibb was the drug provider. The Francis Crick Institute, which receives its core funding from CRUK (FC010110), the UK Medical Research Council (FC010110), the Wellcome Trust (FC010110). For the purpose of Open Access, the author has applied a CC BY public copyright licence to any Author Accepted Manuscript version arising from this submission. TRACERx Renal is funded by NIHR BRC at the Royal Marsden Hospital and Institute of Cancer Research (A109).

## Author Contributions

Conceptualisation, J.L, C.S., S.T. and S.A.Q.; Trial conduct, J.L., S.T., L.P. and L.A. Methodology, S.T., S.A.Q., B.C., T.M., M.M., E.H., G.K., K.L., D.S., L.A. and R.T.; Formal Analysis, M.M., E.H., G.K., D.S., R.T., L.A., K.L. and G.B.; Investigation, A.R., F.B., E.H., I.U., L.A. and M.M; Resources, S.T., J.L., C.S., S.A.Q., M.J-H., A.R., L.A., E.H., F.B., N.F., S.Hazell., D.N. K.J., I.U., P.B., M.S.W., A.A., I.P., W.Y., T.L., K.D. and M.D.V.; Data Curation: L.A., R.M.; Data contribution and interpretation: all; Writing - original draft, L.A., E.H. and M.M.; Writing - review and editing, L.A., S.T., E.H., and M.M. ; Visualisation, M.M., E.H., G.K., L.A., G.B. and R.T.; Supervision, S.T., S.A.Q., B.C., T.M., J.L., C.S.; Project Administration, S.T., J.L., L.A., E.H. and S.A.Q.; Funding Acquisition, J.L, C.S., S.T. and S.A.Q.

## Declaration of Interests

L.A. is funded by the Royal Marsden Cancer Charity. J.A. is a full-time employee of Hoffmann-La Roche AG (Basel, Switzerland). L.P. has received research funding from Pierre Fabre, and honoria from Pfizer, Ipsen, Bristol-Myers Squibb, and EUSA Pharma. S.T. is funded by Cancer Research UK (C50947/A18176), the National Institute for Health Research (NIHR) Biomedical Research Centre at the Royal Marsden Hospital and Institute of Cancer Research (A109), the Kidney and Melanoma Cancer Fund of The Royal Marsden Cancer Charity, The Rosetrees Trust (A2204), Ventana Medical Systems Inc (10467/10530), the National Institute of Health (US) and the Melanoma Research Alliance. R.S. has received non-financial support from Merck and Bristol Myers Squibb; research support from Merck, Puma Biotechnology, and Roche; and advisory board fees for Bristol Myers Squibb; and personal fees from Roche for an advisory board related to a trial-research project; all related to breast cancer research projects. R.S. reports no conflict of interests related to this project. G.K. receives core funding from the Francis Crick Institute (FC0010099). M.J.H. is a Cancer Research UK (CRUK) Clinician Scientist (RCCFEL\100099) and has received funding from CRUK, National Institute for Health Research, Rosetrees Trust, UKI NETs and NIHR University College London Hospitals Biomedical Research Centre. M.J.H. is a member of the Scientific Advisory Board and Steering Committee for Achilles Therapeutics. J.L. has received research funding from Bristol-Myers Squibb, Merck, Novartis, Pfizer, Achilles Therapeutics, Roche, Nektar Therapeutics, Covance, Immunocore, Pharmacyclics, and Aveo, and served as a consultant to Achilles, AstraZeneca, Boston Biomedical, Bristol-Myers Squibb, Eisai, EUSA Pharma, GlaxoSmithKline, Ipsen, Imugene, Incyte, iOnctura, Kymab, Merck Serono, Nektar, Novartis, Pierre Fabre, Pfizer, Roche Genentech, Secarna, and Vitaccess. C.S. acknowledges grant support from Pfizer, AstraZeneca, Bristol-Myers Squibb, Roche-Ventana, Boehringer-Ingelheim, Archer Dx Inc (collaboration in minimal residual disease sequencing technologies) and Ono Pharmaceutical, is an AstraZeneca Advisory Board member and Chief Investigator for the MeRmaiD1 clinical trial, has consulted for Pfizer, Novartis, GlaxoSmithKline, MSD, Bristol-Myers Squibb, Celgene, AstraZeneca, Illumina, Genentech, Roche-Ventana, GRAIL, Medicxi, Bicycle Therapeutics, and the Sarah Cannon Research Institute, has stock options in Apogen Biotechnologies, Epic Bioscience, GRAIL, and has stock options and is co-founder of Achilles Therapeutics. Patents: C.S. holds European patents relating to assay technology to detect tumour recurrence (PCT/GB2017/053289); to targeting neoantigens (PCT/EP2016/059401), identifying patent response to immune checkpoint blockade (PCT/EP2016/071471), determining HLA LOH (PCT/GB2018/052004), predicting survival rates of patients with cancer (PCT/GB2020/050221), identifying patients who respond to cancer treatment (PCT/GB2018/051912), a US patent relating to detecting tumour mutations (PCT/US2017/28013) and both a European and US patent related to identifying insertion/deletion mutation targets (PCT/GB2018/051892). C.S. is Royal Society Napier Research Professor (RP150154). His work is supported by the Francis Crick Institute, which receives its core funding from Cancer Research UK (FC001169), the UK Medical Research Council (FC001169), and the Wellcome Trust (FC001169). C.S. is funded by Cancer Research UK (TRACERx, PEACE and CRUK Cancer Immunotherapy Catalyst Network), Cancer Research UK Lung Cancer Centre of Excellence, the Rosetrees Trust, Butterfield and Stoneygate Trusts, NovoNordisk Foundation (ID16584), Royal Society Research Professorship Enhancement Award (RP/EA/180007), the NIHR BRC at University College London Hospitals, the CRUK-UCL Centre, Experimental Cancer Medicine Centre and the Breast Cancer Research Foundation, USA (BCRF). His research is supported by a Stand Up To Cancer-LUNGevity-American Lung Association Lung Cancer Interception Dream Team Translational Research Grant (SU2C-AACR-DT23-17). Stand Up To Cancer is a program of the Entertainment Industry Foundation. Research grants are administered by the American Association for Cancer Research, the Scientific Partner of SU2C. C.S. also receives funding from the European Research Council (ERC) under the European Union’s Seventh Framework Programme (FP7/2007-2013) Consolidator Grant (FP7-THESEUS-617844), European Commission ITN (FP7-PloidyNet 607722), an ERC Advanced Grant (PROTEUS) from the European Research Council under the European Union’s Horizon 2020 research and innovation programme (835297) and Chromavision from the European Union’s Horizon 2020 research and innovation programme (665233). ST has received speaking fees from Roche, Astra Zeneca, Novartis and Ipsen. ST has the following patents filed: Indel mutations as a therapeutic target and predictive biomarker PCTGB2018/051892 and PCTGB2018/051893 and Clear Cell Renal Cell Carcinoma Biomarkers P113326GB.

### The TRACERx Renal Consortium

The members of TRACERx Renal Consortium are: Lewis Au, Ben Challacombe, Ashish Chandra, Simon Chowdhury, William Drake, Archana Fernando, Nicos Fotiadis, Andrew Furness, Emine Hatipoglu, Karen Harrison-Phipps, Steve Hazell, Peter Hill, Catherine Horsfield, James Larkin, Jose I. Lopez, Teresa Marafioti, David Nicol, Tim O’Brien, Jonathon Olsburgh, Lisa Pickering, Alexander Polson, Sergio Quezada, Sarah Rudman, Scott Shepherd, Charles Swanton, Samra Turajlic, Mary Varia, Hema Verma.

### The PEACE Consortium

The members of PEACE Consortium are: Chris Abbosh, Kai-Keen Shiu, John Bridgewater, Daniel Hochhauser, Martin Forster, Siow-Ming Lee, Tanya Ahmad, Dionysis Papadatos-Pastos, Sam Janes, Peter Van Loo, Katey Enfield, Nicholas McGranahan, Ariana Huebner, Sergio Quezada, Stephan Beck, Peter Parker, Henning Walczak, Tariq Enver, Rob Hynds, Mary Falzon, Ian Proctor, Ron Sinclair, Chi-wah Lok, Zoe Rhodes, David Moore, Teresa Marafioti, Elaine Borg, Miriam Mitchison, Reena Khiroya, Giorgia Trevisan, Peter Ellery, Mark Linch, Sebastian Brandner, Crispin Hiley, Selvaraju Veeriah, Maryam Razaq, Heather Shaw, Gert Attard, Mita Afroza Akther, Cristina Naceur-Lombardelli, Lizi Manzano, Maise Al-Bakir, Simranpreet Summan, Nnenna Kanu, Sophie Ward, Uzma Asghar, Emilia Lim, Faye Gishen, Adrian Tookman, Paddy Stone, Caroline Stirling, Lewis Au, Andrew Furness, Kim Edmonds, Nikki Hunter, Sarah Sarker, Sarah Vaughan, Mary Mangwende, Karla Lingard, Lavinia Spain, Scott Shepherd, Haixi Yan, Ben Shum, Eleanor Carlyle, Steve Hazell, Annika Fendler, Fiona Byrne, Nadia Yousaf, Sanjay Popat, Olivia Curtis, Gordon Stamp, Antonia Toncheva, Emma Nye, Aida Murra, Justine Korteweg, Nahid Sheikh, Debra Josephs, Ashish Chandra, James Spicer, Ula Mahadeva, Anna Green, Ruby Stewart, Lara-Rose Iredale, Tina Mackay, Ben Deakin, Debra Enting, Sarah Rudman, Sharmistha Ghosh, Lena Karapagniotou, Elias Pintus, Andrew Tutt, Sarah Howlett, Vasiliki Michalarea, James Brenton, Carlos Caldas, Rebecca Fitzgerald, Merche Jimenez-Linan, Elena Provenzano, Alison Cluroe, Grant Stewart, Colin Watts, Richard Gilbertson, Ultan McDermott, Simon Tavare, Emma Beddowes, Patricia Roxburgh, Andrew Biankin, Anthony Chalmers, Sioban Fraser, Karin Oien, Andrew Kidd, Kevin Blyth, Matt Krebs, Fiona Blackhall, Yvonne Summers, Caroline Dive, Richard Marais, Fabio Gomes, Mat Carter, Jo Dransfield, John Le Quesne, Dean Fennell, Jacqui Shaw, Babu Naidu, Shobhit Baijal, Bruce Tanchel, Gerald Langman, Andrew Robinson, Martin Collard, Peter Cockcroft, Charlotte Ferris, Hollie Bancroft, Amy Kerr, Gary Middleton, Joanne Webb, Salma Kadiri, Peter Colloby, Bernard Olisemeke, Rodelaine Wilson, Ian Tomlinson, Sanjay Jogai, Christian Ottensmeier, David Harrison, Massimo Loda, Adrienne Flanagan, Mairead McKenzie, Allan Hackshaw, Jonathan Ledermann, Kitty Chan, Abby Sharp, Laura Farrelly, and Hayley Bridger.

## STAR METHODS

Key resources table in separate files

## RESOURCE AVAILABILITY

### Lead Contact

Further information and requests for resources and reagents should be directed to and will be fulfilled by the Lead Contact, Samra Turajlic.

### Materials Availability

This⍰ study did not generate new unique reagents.

### Data and code availability

## EXPERIMENTAL MODEL AND SUBJECT DETAILS

### Clinical studies

ADAPTeR (NCT02446860) is a single-arm, open-label, phase 2 study of nivolumab therapy as pre-operative therapy in metastatic ccRCC. Planned interim analysis took place after six months after the last patient enrolled had their first Response Evaluation Criteria in Solid Tumours (RECIST version 1.1) defined objective response assessment. ADAPTeR was initially approved by NRES Committee London Fulham on 01/12/2014. ADAPTeR is performed in accordance with the ethical principles in the Declaration of Helsinki, Good Clinical Practice and applicable regulatory requirements.

Nivolumab was administered at a dose of 3mg per kilogram of body weight as a 60-minute intravenous infusion every 2-weeks. Eligible patients were 18 years of age or older, had histologic confirmation of advanced or metastatic renal-cell carcinoma (RCC) with predominantly clear cell component with at least one site of disease outside the kidney measurable according to the RECIST version 1.1, with no prior systemic therapy for ccRCC. All patients had an Eastern Cooperative Oncology Group (ECOG) performance status of 0 or 1. Key exclusion criteria were need for immediate nephrectomy, any active, known or suspected autoimmune disease or another condition requiring systemic treatment with either corticosteroids (>10mg daily prednisolone equivalent) or other immunosuppressive medications within 14-days of study drug administration (excluding vitiligo, Type 1 diabetes mellitus, residual hypothyroidism due to autoimmune condition only requiring hormone replacement, psoriasis not requiring systemic treatment or conditions not expected to recur in the absence of an external trigger). During the course of the study, inclusion expanded to those who have had a prior nephrectomy but are suitable for on treatment biopsies. The prognostic factors assessed for the risk categorisation are as per the published IMDC criteria^37^: time to systemic therapy (<1 year), performance status, anaemia, hypercalcaemia, neutrophilia and thrombocytosis. Presence of zero (favourable-risk), one (intermediate-risk), and two or three (poor-risk) factors provides the categorisation.

The primary endpoint was the safety profile of nivolumab given pre- and post-operatively to patients with metastatic ccRCC undergoing nephrectomy. Secondary endpoints were overall response rate (ORR), progression free survival (PFS), and overall survival (OS). Exploratory endpoints pertain to biomarker analyses. Patients deemed clinically suitable for nephrectomy at baseline were scheduled for surgery after the fourth cycle of treatment. Patients not deemed clinically suitable for nephrectomy at baseline would undergo surgery if an excellent clinical response is observed and if surgery was clinically appropriate. Nivolumab treatment was recommenced post-operatively upon sufficient recovery, and until disease progression. Patients who remained clinically unsuitable for nephrectomy continued nivolumab treatment until disease progression.

For translational study sample collection, baseline tumour biopsy via appropriate guidance (ultrasound or computer tomography [CT]) at least 3 days and up to 14 days prior to starting nivolumab was obtained. Tumour multiple regions of nephrectomy specimen were sampled, as well as image guided biopsy of regressing lesions or at disease progression either at site of progression or, if not possible, percutaneous primary renal tumour biopsy, prior to commencement of any subsequent treatment. Blood samples were collected at each tumour sampling timepoint.

Autopsy samples from ADR001, ADR005, and ADR015 were obtained through the PEACE Study (NIHR 18422; NCT03004755), where samples are harvested within ∼48 hours from death. All patients were co-recruited to the TRACERx Renal study (NCT03226886; see secondary author list for the full list of TRACERx Renal consortium investigators). Patient and sample metadata (i.e. age a diagnosis, sex, clinical response, biopsy site) are provided as Table S1 and Supplemental Data Table 1. All the patients provided written informed consent. The protocols, amendments and informed consent forms were approved by the institutional review board or independent ethics committee at each trial site for each trial.

## METHOD DETAILS

### Sample collection

Tumour and normal tissue were collected via image-guided percutaneous biopsies, ex vivo sampling at nephrectomy, and at autopsy. Multiregion samples were obtained with all modalities. For samples obtained at nephrectomy, resected specimens were reviewed macroscopically by a pathologist to guide multiregion sampling for this study and to avoid compromising diagnostic requirements. Spatially separated regions sampled from the ‘‘tumour slice’’ using a 6mm punch biopsy needle. The punch was changed between samples to avoid contamination. The total number of samples obtained reflects the tumour size with a minimum of 3 biopsies that are non-overlapping and equally spaced. Areas which are obviously fibrotic or haemorrhagic are avoided during sampling and every attempt is made to reflect macroscopically heterogeneous tumour areas. Primary tumour regions are labelled as R1, R2, R3.Rn and locations are recorded. Normal kidney tissue was sampled from areas distant to the primary tumour and labelled N1. For all samples collected, each were split into two for snap freezing and formalin fixing respectively, such that the fresh frozen sample has its mirror image in the formalin-fixed sample which is subsequently paraffin embedded. Fresh samples were placed in a 1.8 ml cryotube and immediately snap frozen in liquid nitrogen for >30 seconds and transferred to −80 C for storage. Peripheral blood was collected at the time of surgery and processed to separate buffy coat and peripheral blood mononuclear cells (PBMCs).

### Nucleic acid extraction, DNA and RNA library preparation and sequencing

DNA and RNA were co-extracted from fresh-frozen tumour tissue using AllPrep DNA/RNA mini kit (Qiagen). RNA from peripheral blood mononuclear cells (PBMC) were extracted from blood stored in Tempus tubes using the Tempus™ Spin RNA Isolation Kit (Invitrogen). Germline DNA was isolated from whole blood using the DNeasy Blood and Tissue kit (Qiagen). DNA yield and quality were assessed on TapeStation4200 (Agilent) and Qubit Fluorometric quantification (ThermoFisher Scientific). Samples were normalised to either 3 ug or 200ng and sheared to 150-200bp using a Covaris-E220 or LE220-plus. Agilent SureSelectXT enriched libraries were constructed following the manufacturer’s manual or automated (using the Agilent Bravo liquid handling platform) SureSelectXT Target Enrichment System for Illumina Paired-end Multiplexed Sequencing Library protocol. Hybridisation and capture were performed using the Agilent SureSelectXT Human All Exon v5 capture library. Final libraries were sequenced to a target coverage of 250x with 101bp paired-end reads multiplexed on the Illumina HiSeq4000 sequencing platform. The extracted RNA was normalised to 100ng for library construction using RNA-Ribozero (ribodeplete) Library Preparation Kits. The prepared libraries were multiplexed and QC’ed before paired-end sequencing with target coverage of 50 million reads per sample on HiSeq4000 sequencing platforms (Illumina). RNA was extracted from blood for TCR sequencing from the following cases and timepoints: all cases (n=15) pre- and post-treatment.

### SNV, and INDEL calling from multiregion WE sequencing

Paired-end reads (2×100bp) in FastQ format sequenced by Hiseq were aligned to the reference human genome (build hg19), using the Burrows-Wheeler Aligner (BWA) v0.7.15. with seed recurrences (-c flag) set to 10000 ^90^. Intermediate processing of Sam/Bam files was performed using Samtools v1.3.1 and deduplication was performed using Picard 1.81 (http://broadinstitute.github.io/picard/).Single Nucleotide Variant (SNV) calling was performed using Mutect v1.1.7 and small scale insertion-and-deletions (INDELs) were called running VarScan v2.4.1 in somatic mode with a minimum variant frequency (--min-var-freq) of 0.005, a tumour purity estimate (--tumour-purity) of 0.75 and then validated using Scalpel v0.5.3 (scalpel-discovery in --somatic mode) (intersection between two callers taken) ^91-93^. SNVs called by Mutect were further filtered using the following criteria: i) ≤5 alternative reads supporting the variant and variant allele frequency (VAF) of ≤1% in the corresponding germline sample, ii) variants falling into mitochondrial chromosome, haplotype chromosome, HLA genes or any intergenic region were not considered, iii) presence of both forward and reverse strand reads supporting the variant, iv) >5 reads supporting the variant in at least one sample, v) variants were required to have a VAF of 0.01 in at least one sample, vi) sequencing depth need to be ≥20 and ≤3000 across all samples. Dinucleotide substitutions (DNV) were identified when two adjacent SNVs were called and their VAFs were consistently balanced (based on proportion test, *P*≥0.05). In such cases the start and stop positions were corrected to represent a DNV and frequency related values were recalculated to represent the mean of the SNVs. Variants were annotated using Annovar^94^. Individual tumour biopsy regions were judged to have failed quality control and excluded from analysis based on the following criteria: i) sequencing coverage depth below 100X, ii) low tumour purity such that copy number calling failed. Driver variants are manually reviewed and predicted for variant effect, and variant annotations on the heatmap are only for confident driver events.

### Methylation specific PCR

Methylation of the *VHL* promoter was detected after bisulphite treatment of 500ng of patient DNA using the EZ DNA Methylation-Direct kit (Zymo Research). Bisulphite treated DNA was amplified in the PCR using methylation specific oligonucleotides followed by Big Dye terminator Sanger sequencing. Methylation was confirmed by comparing and contrasting patient tumour and normal renal tissue for methylation protected CpG sequences.

### Neoantigen calling

Neoantigen predictions were derived by first determining the 4-digit HLA type for each patient, along with mutations in class I HLA genes, using POLYSOLVER^95^. Next, all possible 9, 10 and 11-mer mutant peptides were computed, based on the detected somatic non-synonymous SNV and INDEL mutations in each sample. Binding affinities of mutant and corresponding wildtype peptides, relevant to the corresponding POLYSOLVER-inferred HLA alleles, were predicted using NetMHCpan (v3.0)^96^ and NetMHC (v4.0)^97^. Neoantigen binders were defined as strong binders if their %rank was below <0.5 for the mutant and >0.5 for the wildtype protein.

### TMB, fsINDEL burden, neonatigen burden, wGII

To calculate sample-level TMB, fsINDEL burden and neoantigen burden, variants were filtered with the same criteria as described above for multi-region WE sequencing but on a per sample-basis. Variants were restricted to positions falling inside the targeted capture range (±50bp padding). Tumour mutational burden (TMB) was calculated as the number of exonic non-synonymous SNVs per mega base. The frameshift INDEL (fsINDEL) burden was calculated as the total number of exonic frameshift INDELs per sample. The neoantigen burden was calculated as the total number of predicted strong binders per sample. The average proportion of the genome with aberrant copy number, weighted on each of the 22 autosomal chromosomes, was estimated as the weighted genome instability index (wGII).

### SNP calling

Single nucleotide polymorphisms (SNPs) were called in the germline sample using Platypus v0.8.1 with default parameters apart from --genIndels=0 and --minMapQual=40. Tumour regions were genotyped based on the variants identified in the germline (parameters set to --minPosterior=0 -- getVariantsFromBAMs=0). SNPs with a minimum coverage of 50x in the germline and the tumour sample were used for allele-specific copy number segmentation.

### Copy number analysis

CNVkit v0.7.3 was used with default parameters on paired tumour-normal sequencing data^98^. Outliers of the derived log2-ratio (logR) calls from CNVkit were detected and modified using Median Absolute Deviation Winsorization before case-specific joint segmentation of fresh-frozen samples to identify genomic segments of constant logR^99^. Formalin-fixed and paraffin-embedded (FFPE) samples were segmented separately while leveraging the segment information from the fresh-frozen samples. Copy number alterations were called as losses or gains relative to overall sample wide estimated ploidy. Driver copy number was identified by overlapping the called somatic copy number segments with putative driver copy number regions identified by Beroukhim et al.^100^. Allele-specific segmentation was performed using the paired PSCBS method after removal of single-locus outliers (R package PSCBS v0.61.0 ^101^).

### Purity and ploidy estimate

Tumour sample purity, average ploidy and absolute allelic copy number per segment were estimated using ABSOLUTE v1.2 in allelic mode^102^. In line with recommended best practice all ABSOLUTE solutions were reviewed by 3 researchers, with solutions selected based on majority vote. Purity assigned 0.1 for samples below ABSOLUTE estimate thresholds for comparison analysis of samples between responders and non-responders.

### Subclonal deconstruction

To estimate the CCF of a mutation, we used the following formula:

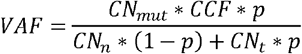

Where VAF is the variant allele frequency of the mutation, p the estimated tumour purity, CN_mut_ the number of copies carrying the mutation and CN_t_ the local copy number in the tumour cells. CN_n_ is the local copy number in the non-tumour proportion of the sample which was assumed to be 2. The CN_mut_ and CCF were estimated through iteration of all possible combinations of CCF (range 0.01 to 1, by 0.01) and CN_mut_ (range 1 to CN_t_, by 1) using the formula above to identify the best fit CCF.

### Genomic contraction, persistence and expansion analysis

For each patient with matched pre- and post-treatment WES data (N=8 patients), the pre-treatment CCF was compared to the CCF in the post-treatment samples. In patients with multiple pre-treatment samples, the median pre-treatment CCF was used. In each post-treatment sample, all nonsynonymous SNVs and fsINDELs were assigned to one of the following categories: “Genomic contraction”, “Genomic persistence”, “Genomic expansion”^39^. A mutation was defined to have undergone genomic contraction if the CCF decreased by ≥10% from pre- to post-treatment or if the mutation was present in the pre-treatment but not the post-treatment sample; genomic expansion if the CCF increased by ≥10% from pre- to post-treatment or if the mutation was present in the post-treatment but not the pre-treatment sample; genomic persistence if the CCF in the post-treatment sample was within the range of ±10% of the pre-treatment CCF. The proportion of mutations falling into each category was calculated for nonsynonymous SNVs and fsINDELs and repeated with only neoantigen encoding mutations. An enrichment test (Fisher’s exact test) was performed to determine whether mutations which encode neoantigens were more likely to undergo genomic contraction than the remaining nonsynonymous SNVs and fsINDELs.

### Mutational signature analysis

Mutational signatures were estimated using the deconstructSigs package in R^103^. Sample specific mutational signature analysis was restricted to samples with at least 50 mutations.

### Analysis for mismatch repair deficiency

Analysis for mutations in the following nominated genes was performed: *POLD3, MLH3, MSH6, RPA4, LIG1, MLH1, MSH2, MSH3, PCNA, PMS2, POLD1, POLD2, POLD4, RFC1, RFC2, RFC3, RFC4, RFC5, RPA1, RPA2, RPA3, SSBP1, EXO1*.

### Analysis for mutations associated with defective antigen presentation

Analysis for mutations in the following nominated genes was performed: *B2M, CIITA, IRF1, PSME1, PSME2, PSME3, ERAP1, ERAP2, HSPA, PSMA7, HSPC, HSPBP1, TAP1, TAP2, TAPBP, CALR, CNX, CANX, PDIA3*.

### Detection of *B2M* mutations by Sanger sequencing

Validation of the *B2M* mutation was performed using PCR followed by Big Dye Terminator Sanger sequencing on the ABI 3700. 20ng of patient DNA was amplified for exon 1 of *B2M*, to enable detection of *B2M*:c.42_45delTCTT:p.S14fs. PCR conditions involved 35 cycles of denaturation at 950C, followed by oligonucleotide primer annealing at 55oC and sequence extension at 720C using Qiagen Taq polymerase and reagents. Oligonucleotide sequences used are: Forward: aacgggaaagtccctctctc; Reverse: agatccagccctggactagc.

### Bulk RNAseq data processing

RNAseq data were mapped to the hg19 reference human genome using the STAR^104^ algorithm, and transcript and gene abundance were estimated by RSEM^105^ with default parameters. Samples were excluded if they had less than 15,000 genes detected.

### Human endogenous retrovirus (HERV) analysis

Expression of previously annotated HERVs^28, 50, 51^ was analysed. HERV loci used in these three studies^28, 50, 51^ were taken from Mayer et al. ^53^ and Vargiu et al.^54^ with 66 and 3173 loci respectively. BLASTn was used to match example sequences from HERVs in Mayer et al. to GRCh38, chromosome coordinates with the greatest homology over the greatest length were taken as the best match. The Lift Genome Annotations tool from UCSC (https://genome.ucsc.edu/cgi-bin/hgLiftOver) was used to convert annotated GRCh37 HERV loci coordinates from Vargui et al. to GRCh38 coordinates. Comparing the new coordinates, 47 of the 66 HERVs from Mayer et al. were present in the list of 3173. Coordinates of all the unique elements were then compared to a custom repeat region annotation previously built using the Dfam 2.0 library (v150923) for GRCh38^57^. For this custom annotation, different regions of the same provirus (e.g. the LTR and internal genes) were annotated separately, these regions were merged to allow accurate quantitation of reads from the same provirus^57^. LTR-containing repeat regions from the custom annotation had to begin, end, or be fully contained within previously annotated loci to be considered a match, a buffer of 5 bases either end of the locus was included. Previously annotated HERV loci from Mayer et al. and Vargiu et al. were found to overlap multiple repeat regions per locus in our custom annotations, or were found to overlap no repeat regions at all. Some loci also overlapped other endogenous retroelement types such as LINEs and SINEs, as well as overlapping canonical gene exons. For this analysis, only expression of matching LTR-containing elements was considered rather than expression of all repeats and genes overlapping previously annotated loci. Expression was measured using read counts calculated by the featureCounts function from the Subread package^106^ (with parameters -p -C -B -f -T 2), multi-mapping reads were not counted. Analysis for purified immune cell subset expression were performed on publicly available datasets from Linsley et al. ^107^ (accession no. GSE60424 (GEO)) and Kazachenka et al.^108^ (E-MTAB-8208 (EMBL-EBI)). LTR-overlapping transcripts expressed highly specifically in ccRCC were previously described^57^. These transcripts were identified through de novo transcriptome assembly and their expression quantified in by transcript per million calculations, as previously described^57^.

### Differential gene expression analysis, pathway analysis and gene set enrichment

DESeq2^109^ was used for differential expression analysis, using the binomial Wald test after estimation of size factors and estimation of dispersion. To identify genes differentially expressed between responders and non-responders, we considered only transcripts with normalized count number >5 in at least 5 patients. Pathway analysis was performed using the R package XGR^108^ using the gene ontology biological process (GOBP) databases. Induced and suppressed transcripts were analysed separately against the background of all tested transcripts. The “lea” ontology algorithm was used.

### T-cell subset gene signature

Gene signature or single gene enrichment was evaluated using RSEM abundance, z score scaled across all samples for which RNA-Seq was available. Signature analysis was performed using 22 immune-related signatures listed below: i) the Danaher immune score is a 60-marker gene signature derived from pan-cancer RNAseq analysis for 14 immune cell populations, where marker genes have been benchmarked against histological tumour-infiltrating lymphocyte (TIL) estimates and flow cytometry data^58, 111^; ii) IMmotion150 ^20^; iii) Javelin101^21^.

1. Danaher Tcells : CD3D, CD3E, CD3G, CD6, SH2D1A, TRAT1
2. Danaher CD8 : CD8A, CD8B
3. Danaher Cytotoxic : CTSW, GNLY, GZMA, GZMB, GZMH, KLRB1, KLRD1, KLRK1, PRF1, NKG7
4. Danaher Bcells : BLK, CD19, MS4A1, TNFRSF17, FCRL2, KIAA0125, PNOC, SPIB, TCL1A
5. Danaher NKcells : NCR1, XCL2, XCL1
6. Danaher CD45 : PTPRC
7. Danaher DC : CCL13, CD209, HSD11B1
8. Danaher CD8Ex : CD244, EOMES, LAG3, PTGER4
9. Danaher Mac : CD163, CD68, CD84, MS4A4A
10. Danaher Mast : MS4A2,TPSAB1,CPA3,HDC,TPSB2
11. Danaher Neut : CSF3R, S100A12, CEACAM3, FCAR, FCGR3B, FPR1, SIGLEC5
12. Danaher NKCD56 : IL21R, KIR2DL3, KIR3DL1, KIR3DL2
13. Danaher Th1 : TBX21
14. Danaher Treg : FOXP3
15. IMmotion150 Angio : VEGFA, KDR, ESM1, PECAM1, ANGPTL4, CD34
16. IMmotion150 Teff : CD8A, IFNG, PRF1, EOMES, CD274
17. IMmotion150 Myeloid : CXCL1, CXCL2, CXCL3, CXCL8, IL6, PTGS2
18. Javelin101 TCR : CD3G, CD3E, CD8B, THEMIS, TRAT1, GRAP2, CD247
19. Javelin101 TCell : CD2, CD96, PRF1, CD6, IL7R, ITK, GPR18, EOMES, SIT1, NLRC3
20. Javelin101 NK : CD2, CD96, PRF1, CD244, KLRD1, SH2D1A
21. Javelin101 chemo : CCL5, XCL2
22. Javelin101 other : CST7, GFI1, KCNA3, PSTPIP1

The signature score was calculated as the arithmetic mean of z score scaled expression of all genes in that signature for each sample.

### TCR sequencing

TCR β-chain sequencing was performed by utilizing whole RNA extracted from tissue samples or from cryopreserved PBMC samples, by using a quantitative experimental and computational TCR sequencing pipeline described previously^112-115^. An important feature of this protocol is the incorporation of a UMI attached to each cDNA TCR molecule that enables correction for PCR and sequencing errors, which allows higher quantitative precision compared to alternate protocols in the TCR sequences retrieved^113, 116^. The suite of tools used for TCR identification, error correction and CDR3 extraction is freely available at https://github.com/innate2adaptive/Decombinator.

For each TCR, we computed the abundance as the count of UMIs mapping to this TCR divided by the total number of UMIs in the sample. If several samples were available at a given patient-timepoint pair, the resulting abundance was calculated as the sum of counts for this TCR across the available samples divided by the sum of total counts across these samples.

### Repertoire similarity measure

The similarity between two TCR repertoires was assessed with the normalised dot product (also known as the cosine similarity) between the vectors of TCR abundance. This measure is a well-established metric widely used in machine learning to compare numerical vectors and gives a value between 0 (no similarity, that is, orthogonal vectors) and 1 (complete similarity, from vectors with an identical magnitude and direction in the feature space). Each pair of repertoires is represented as two vectors of equal length, indexed by the union of TCRs found in both repertoires and containing the number of times each TCR is detected in each of the two repertoires (each position contains an integer ≥0). The similarity between the two vectors is given as

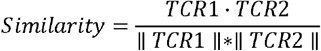

where and are the abundance vectors, represents the vector product and paired vertical bars represent the Euclidean norm of the vector.

For longitudinal similarity (Figures 4, S14, and S15), the similarity measure was performed on the TCR abundance vectors derived from (patient, timepoint) pairs.

For spatial similarity (Figure S13), the similarity measure was performed on the TCR abundance vectors derived from each sample within a (patient, timepoint) pair. For this analysis, samples from different timepoints were not compared.

### Repertoire clonality index

The clonality index was estimated for each sample by using the command entropy from the entropy R package, on the basis of the observed frequency of the TCRs in that sample

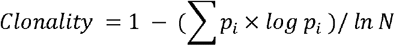

where *p*_*i*_ is the frequency of the ith TCR in the repertoire and is the number of TCRs in that repertoire.

### Classification of expanded, contracted and persistent TCRs

The difference in abundance between Pre-treatment and On-treatment was calculated with the poisson.test function in R, as the data were counts. TCRs with *P* values above 0.01 were labelled as persistent.

### Classification of expanded TCRs

We counted the number of TCRs detected with frequencies above a range of frequency thresholds in the tumour repertoires. To measure how such defined expanded TCRs were representative of the shape of the TCR distribution captured by the clonality score, we computed the prevalence of the expanded population amongst the entire repertoire, for each threshold. To do so, we took the sum of counts for expanded TCRs and divided it by the sum of all counts in the sample. The proportion obtained was then correlated to the matched clonality score with the Spearman’s rank correlation.

To focus on the most expanded TCRs (Figures 4E-4F and S15-S16), we examined those present above a threshold frequency of 2/1,000 (corresponding to the top 1% of the empirical TCR frequency distribution). At this threshold, which we already described in previously published work ^61^, the correlation between clonality and proportion of repertoire occupied by expanded TCRs is very strong and the number of TCRs labelled as expanded is greater than for higher thresholds for which this correlation is also significant, which enables to keep the greatest amount of data whilst still applying a stringent filtering step.

### CDR3 amino acid clustering

The pairwise similarity between pairs of TCRs was measured on the basis of amino acid triplet sharing. Sharing was quantified using the normalized string kernel function stringdot (with parameters stringdot (type = ‘spectrum’, length = 3, normalized = TRUE) from the Kernlab package. The kernel is calculated as the number of amino acid triplets (sets of three consecutive amino acids) shared by two CDR3s, normalized by the number of triplets in each CDR3 being compared. The TCR similarity matrix was converted into a network diagram by using the iGraph package in R. Two TCRs were considered connected if the similarity index was >0.82 (threshold previously optimised in a separate study).

Per (patient, timepoint) pair, we counted the number of clusters containing an expanded CDR3. To normalize the counts of clusters obtained (*Nreal*)for the input size, for each sample, we randomly selected, outside of the real clustering structure, a number of CDR3s equal to the number of expanded CDR3s in that sample and looked for clusters around those. This control step was repeated 10 times for each (patient, timepoint) pair and we computed the average number of clusters obtained for those control (*Ncon*) and used *Nreal*/ *Ncon* as the normalised cluster count value (Figure 4F, Figure S15F).

For Figure S15D, as depicted in Figure S15E, we used the clustering structure built as described above for Pre-treatment samples and retrospectively labelled expanded clones at that time-point as maintained if they were also expanded post-treatment or as replaced if they were not. By doing so, we could derive the number of Pre-treatment clusters containing maintained (resp. replaced) expanded clones which was then divided by the initial count of maintained (resp. replaced) expanded clones present in that sample to obtain the proportion displayed in FigureS15D.

### Frequency ratio

We wanted to capture the rate of clonal replacement that occurs in the tumour repertoires. To do so, for each expanded TCR at baseline that could also be detected after treatment, we computed the ratio of the observed frequency at baseline divided by the observed frequency after-treatment. To derive a metric for each patient, we computed the average of ratio scores obtained for all expanded TCRs at baseline (those that could not be detected after treatment were excluded).

### Multiplex Immunofluorescence Staining and Image Analysis

FFPE blocks were cut in 2 micron thick slides. The slides were baked for 60 minutes and stained using the antibodies listed below and opal fluorophores. Leica Bond III machine was used for the immunofluorescence staining. Images of the stained slides were acquired by using the Vectra 3 automated quantitative pathology imaging system (Akoya Biosciences). Matching H&E image of each slide was reviewed by a pathologist and areas to annotate on the immunofluorescent images for analysis were identified. Necrotic and stromal areas as well as non-tumour areas were excluded and tumour areas were scored. Slides for patient ADR009 were not evaluable due to necrosis. Total of 61 samples (41 pre-treatment and 20 on treatment samples) for the first mIF panel and 60 samples (40 pre-treatment and 20 on treatment samples) were for the second IF panel were used for analysis. The following antibodies were used for mIF staining: CD3 (Mouse monoclonal, LN10, 1:100 dilution on Opal 520 in 1:50 dilution), CD4 (Mouse monoclonal, 4B12, 1:50 dilution on Opal 540 in 1:100 dilution), CD8 (Mouse monoclonal, 4B11, 1:100 dilution on Opal 540 in 1:150 dilution and on Opal 620 in 1:150 dilution), FoxP3 (Mouse Monoclonal, 236A/E7, 1:80 dilution on Opal 570 in 1:150 dilution), CD163 (Mouse monoclonal, 10D6, 1:100 dilution on Opal 690 in 1:50 dilution), Granzyme B (Mouse monoclonal, 11F1, 1:80 dilution, on Opal 620 in 1:150 dilution)

Up to 25 multispectral images (MSI) were acquired per slide depending on the size of the tumour to include all representative areas of the tumour. Representative MSIs from different slides were used while training the algorithms for each marker. Scoring of each slide was performed using the inForm software on Vectra. The quality and accuracy of the scoring was checked by two clinicians one of whom was a histopathologist. MSIs with poor tissue quality were excluded from the analysis. Merged data obtained by using the inForm software was analysed using the phenoptrReports tool (Akoya Biosciences) on R. T-cells subsets (CD8^+^, CD4^+^ effectors, Tregs and CD8^+^CD4^+^ double positive cells) were scored both out of total cells counted on each slide and out of the total T cells counted. CD163 cells were scored out of total cells counted per slide. Overall granzyme expression was scored in relation to the total T-cell and CD163^+^ cell count. Granzyme B expression on CD8^+^ cells was scored out of the total CD8^+^ cells. Median scoring value was used for each patient per time point and two-sided Mann-Whitney U test was used for statistical analysis of the data.

The mIF and mIHC antibody panels were designed to evaluate T-cell subsets, B-cells, myeloid cells, and GZMB expression. This was conducted given 1) double positive (CD8^+^CD4^+^) T-cells with high degrees of TCR clonality have previously been described in ccRCC^117^; 2) myeloid inflammation has been associated with blunting of anti-tumour T-cell activity in metastatic ccRCC^20^; and 3) high tumour infiltration with B-cells and plasma cells have previously been shown to correlate with favourable clinical outcomes across cancer types ^118-120^.

### Immunohistochemistry

Formalin-fixed paraffin-embedded tissue sections of clear-cell renal cell carcinoma and normal tonsil tissues were subjected to haematoxylin and eosin and multiplex immunostaining. The primary antibodies used for multiplex immunolabeling are as follows: CD19 (Rabbit monoclonal, SP291, 1:10000 dilution), CD138 (Mouse monoclonal, MI15, 1:100 dilution), PD-1 (Mouse Monoclonal, NAT105/E3, 1:2 dilution).

To establish optimal staining conditions each antibody was tested and optimized on 2-4 um cut tissue sections of human reactive tonsil and normal kidney by applying conventional single immunohistochemistry. In brief sections were de-waxed and re-hydrated prior to the multiplex immunolabeling whose procedure was adapted and performed according to the established protocol described elsewhere^121^. Total of 59 samples (40 pre-treatment and 19 on treatment samples) for the mIHC panel.

### Staining assessment and data handling

Specificity of the staining was assessed by a haematopathologist with expertise in multiplex-immunostaining. Scanned slide images were obtained with the use of NanoZoomer Digital Pathology System (Hamamatsu, Japan). Total of 60 samples (41 pre-treatment and 19 on treatment samples) were used for analysis.

### Flow cytometry

Renal tumour resections and normal tissue were cut into small pieces (2-3mm) by using sterile disposable scalpel plus forceps in RPMI (Sigma-Aldrich) with Collagenase I (Sigma-Aldrich) (for ADR013 tumour and normal tissue), Liberase (for ADR001 tumour tissue) and DNAse I (Roche) and was digested for 1 hour at room temperature using the gentleMACS dissociator (Miltenyi Biotec). The digest was passed through a 70-µm cell strainer by using 5-10 ml of RPMI containing 2% fetal bovine serum (FBS) to obtain a single cell suspension. Lymphocytes were obtained from the single cell suspension by using Ficoll Paque Plus (GE Healthcare) density gradient centrifugation (750g for 10 min). Isolated lymphocytes were washed with RPMI and 2%FBS and cryopreserved in 90% FBS with 10% dimethyl sulfoxide (Sigma–Aldrich). PBMCs were isolated from blood samples collected in Vacutainer EDTA blood collection tubes (BD) using Ficoll Paque Plus (GE Healthcare) density gradient centrifugation and cryopreserved in in 90% FBS with 10% dimethyl sulfoxide (Sigma–Aldrich).

Thawed lymphocytes were washed with 1x phosphate-buffered saline (PBS) and were stained with the antibodies listed below. Antibody mastermixes were prepared in Brilliant Staining Buffer (BD). eBioscience™ Foxp3 / Transcription Factor Staining Buffer Set was used for the intracellular staining. Samples were stained using the following antibodies: CD8 (RPA-T8, BUV496), CD45RA (HI100, BUV563), CD4 (SK3, BUV615), CD38 (HIT2, BUV737), CD3 (SK7, BUV705), FoxP3 (206D, BV421), CCR4 (L291H4, BV510), Viability dye (Yellow Fluorescent reactive dye, BV570), CD57 (QA17A04, BV605), Ki67(B56, BV650), CD39 (TU66, BV711), CCR7 (G043H7, BV750), CD69(FN50, BV785), CD103 (Ber-ACT8, BB515), CXR5 (RF8B2, PerCp-Cy5-5), TCF-7 (7F11A10, PE), Granzyme B (GB11, PE-CF594), CD25 (M-A251, PE-Cy5), PD-1 (EH12.2H7, PE-Cy7), TOX (REA473, APC), HLA-DR (LN3, AF700), IgG4(Biotin), 4-1BB (4B4-1, BUV661), TIM3 (7D3, BV650), KLRG1 (13F12F2, Superbright702), CD27 (O323, BV750), ICOS (C398.4A, BV785), EOMES (WD1928, Percp-eFlour710), CTLA-4 (L3D10, APC), GITR (108-17, APC/Fire-750), Streptavidin (BUV395). The samples were acquired on the BD Symphony flow cytometer. Data was analysed using the FlowJo (version 10).

### PD-1 competition binding assay to evaluate anti-PD1 monoclonal antibody binding

PBMC isolated from healthy individuals were activated *in vitro* using plate coated anti-CD3 and soluble anti-CD28 with 100IU IL-2 per well. 50ul (5ug/ mL solution) anti-CD3 was used to coat wells of a 96 well plate which was kept at 4°C overnight. Two washes using 200ul of PBS were performed to remove unbound antibodies the next day. Subsequently, 2 × 10^5^ PBMC were added into each well with subsequent addition of soluble anti-CD28 (2ug/ mL). The plate was placed into a humidified 37°C incubator for 72 hours. Following this period, the wells containing activated PBMC were either incubated with 50ul (2.5mg) pembrolizumab or PBS control for 30 minutes. PBS washes were used to remove unbound therapeutic antibodies. Flow cytometric staining of CD3, PD-1 and anti-IgG4 was performed thereafter.

### Single-cell RNA/TCR Sequencing

Tumour infiltrating lymphocytes from ADR001 and AD013 were stained with CD3 (PE, SK7 clone), IgG4 (Biotinylated) and Streptavidin (BV650) antibodies for flow cytometry. Stained cells were FACS sorted as CD3^+^IgG4^-^ (40,000 cells) and CD3^+^IgG4^+^ (20,000 cells) for ADR001 and CD3^+^IgG4^-^ (50,000 cells) and CD3^+^IgG4^+^ (90,000 cells) for ADR013. FACS sorted cells were single cell sorted using the 10X Genomic machine. The sorted cells were processed using the 10X Genomic Chromium Next GEM Single Cell 5’ Reagents Kit V2 (dual index) for 5’gene expression library construction and V(D)J library construction. The samples were sequenced on the NextSeq using the High Output Kit v2.5 (150 Cycles).

FASTQ files containing gene expression (GEX) and VDJ were demultiplexed using cellranger mkfastq (10x Genomics). GEX reads were aligned to GRCh38 and counted using cellranger count, VDJ reads were aligned to cellranger’s GRCh38 VDJ reference dataset using cellranger vdj. Expression matrices were analysed using the Seurat package^122^. To remove technical variation in the data, TCR, ribosomal and heat-shock protein genes were removed from the analysis, also cells with mitochondrial reads making up >10% total read content were removed. 8382 CD3^+^IgG4^-^ and 10083 CD3^+^IgG4^+^ cells in ADR013; and 4648 CD3^+^IgG4^-^ and 3343 CD3^+^IgG4^+^ cells in ADR001 were retained after quality control filtering. Datasets were integrated using SCTransform integration^123^ using the recommended parameters and regressing the % mitochondrial read content. PCA and UMAP dimensional reduction (dims = 1:30) and clustering (res = 0.3) was then performed using RunPCA, RunUMAP, FindNeighbours, and FindClusters. Publicly available gene signatures for T cell states were obtained from the following publications: Schietinger et al.^124^, Thommen et al.^125^, Guo et al.^126^, Li et al. ^127^, Yost et al.^128^, Miller et al.^69^, Zhou et al.^129^, and Litchfield et al.^130^ (**Supplemental Data Table 3**). The proportion of reads mapping to the genes in each signature for each cell was then calculated using PercentageFeatureSet. All differential gene expression analysis were carried out on log normalised gene expression values (using NormalizeData, default parameters) using the MAST algorithm^131^ within FindMarkers. GOBP analysis was carried out using the XGR package^110^ using the “lea” algorithm. scTCR data was analysed using scRepertoire^132^. Cells were considered of the same clone if they contained a matching TRB sequence and CDR3 gene.

## QUANTIFICATION AND STATISTICAL ANALYSIS

Statistical analysis was performed in R and GraphPad Prism 8. Correlation was carried out with the Spearman nonparametric rank correlation test. We used mixed effect modelling when appropriate. We used the Mann– Whitney two-tailed paired or non-paired nonparametric tests (as appropriate) to determine whether two independent samples were selected from the same populations. *P* values were considered significant if less than 0.05, and significance values were corrected for multiple testing by Bonferroni correction when appropriate. High dimensional flow cytometry analysis was performed using FlowJo 10. Analyses and visualization of HERV expression were additionally performed in Qlucore Omics Explorer (Qlucore, Lund, Sweden). Data visualization was performed in BioRender, R and GraphPad Prism 8.

## ADDITIONAL RESOURCES

Clinical trial registry numbers:

ADAPTeR: https://clinicaltrials.gov/ct2/show/NCT02446860

TRACERx Renal: https://clinicaltrials.gov/ct2/show/NCT03226886

PEACE: https://clinicaltrials.gov/ct2/show/NCT03004755

**Supplementary Figure Legends**

**Figure S1. Samples overview, and correlations between nivolumab response and mutational features**

**(A)** Consort diagrams for samples that underwent whole-exome sequencing, RNAseq, TCRseq, and multiplex immunofluorescence or immunohistochemistry analyses. **(B)** Boxplots showing no significant correlations between TMB, fsINDEL load, and wGII to nivolumab response. Median pre-treatment values are shown. **(C)** Genomic contraction analysis results including neoantigens. Two-sided Mann–Whitney test was performed. *P* value >0.05 considered not significant.

**Figure S2. Schematic diagrams of ADR015 and ADR005 showing pre-/post-treatment, post-mortem sampling, and evolution of metastatic disease**

**(A)** Clinical timeline for ADR015, and **(B)** genomic data are shown. **(C)** Clinical timeline for ADR005, and **(D)** PEACE samples with available TCRseq data. **(E)** The proportion of TCRs that were expanded both Pre-treatment and Post-treatment during life (n=5) detected in each post-mortem sample, only samples where the detection rate is greater than 0 are displayed. 3/5 were detected in the lung metastatic and 1/5, 1/5, 2/5 and 3/5 were detected in region 1, region 2, region 3 and region 4 of the primary site, respectively. The median number of TCR sequences retrieved per post-mortem sample was 163 (range: 20-1340), and immune infiltration across sites were uniformly low in all regions, as scored by expert pathologist review of haematoxylin and eosin stain slides (data not shown).

**Figure S3. Correspondence of HERV annotation and expression in immune cell types**

**(A)** Comparison of HERV annotations by Attig et al., Vargiu et al. and Mayer et al. Three examples are shown, depicting the position of GENCODE annotated genes, Dfam 2.0 annotated repeats and a representative RNAseq read pileup. **(B)** Expression of HERVs previously associated with ccRCC or with nivolumab response, in the indicated purified immune cell types from public RNAseq datasets GSE60424 (top) and E-MTAB-8208 (bottom). Note the expression of LTR/ERVK|HERVK9-int∼MER9a1|6|29876165|29881829 integration within the HLA locus in most immune cell subsets and of the LTR/ERV1|LTR7|1|207633751|207634199 integration in neutrophils.

**Figure S4. Violin plots comparing response groups at both timepoints by Danaher, IMMOTION150, Javelin101 signatures and by individual gene expression**.

See **STAR Methods** for details of signature analysis. The two-sided Mann–Whitney test performed on one value per patient (score averaged by median value across biopsies if several available at a given time point), significant *P* value are indicated (*: *P*<0.05; **: *P*<0.01). *R - responders; N-R - non-responders*.

**Figure S5. Immune cells subset comparisons of pre- and post-treatment samples**

**(A)** Immune cell subset expression levels in non-responders and responders on treatment are shown. **(B)** Expression level of T-cell subsets out of total cells counted is shown. **(C)** Ratio of T-cells subsets in non-responders and responders at baseline and on treatment; CD3^+^ T-cells to CD163^+^ myeloid cells, and CD8^+^ T-cells to CD163^+^ myeloid cell ratios between responders and non-responders on treatment is shown. **(D)** Change in total GZMB expression and on CD8^+^ T cells from pre-treatment (six patients) to post-treatment (week-9) is shown (three patients).

**Figure S6. Clonotype dynamics in PBMC and intra- and inter-patient TCR repertoire heterogeneity**

**(A)** The TCR repertoires of multiple biopsies from a patient’s tumour were sequenced and a pairwise comparison of the repertoires of different biopsies from the same timepoint was performed by using the cosine metric (**STAR Methods**). The pairwise intratumoural TCR repertoire similarity is shown for each patient. Each circle represents a comparison between two samples from the same patient (n = 87 total comparisons from 12 patients). Red (resp. blue) circles indicate a pair of biopsies originating from the same site (resp. different metastatic sites). **(B)** Heat maps showing the pairwise similarities of a selection of 5 biopsies in the post-treatment nephrectomy for ADR001 (top) and ADR013 (bottom). Biopsies were selected based on comparable TCR counts. **(C)** Correlated clone sizes in blood samples. Scatter plots of blood clone size after treatment and before treatment are shown for all patients. Clones are colored by expansion/contraction status (**STAR Methods**). **(D)** The peripheral TCR repertoire clonality score pre-treatment and on-treatment is shown for each patient. Patients are split between responders and non-responders. Mixed-effect model *P* value shown. **(E)** The number of intratumoural (left panel) and peripheral (right panel) clones labelled as expanded or contracted between timepoints, per patient, normalized for the total number of clones tested. Two-sided Mann–Whitney tests *P* value shown. **(F)** The peripheral cosine score between pre-treatment and on-treatment is shown for each patient. Patients are split between responders and non-responders. Two-sided Mann–Whitney test *P* value shown; n=12 patients.

**Figure S7. Additional expanded TCRs metrics**

**(A)** The arithmetic mean of Pre/Post frequency ratios of clones expanded pre-treatment, per patient. Two-sided Mann–Whitney test *P* value shown. **(B)** The Spearman’s rank correlation coefficient and *P* value (shown above each point; n=14 patients) for the relationship between the clonality score and the proportion of the intratumoural repertoire pre-treatment occupied by expanded clones defined by different frequency thresholds (ranging from all TCRs (threshold of zero) up to those found at a frequency of ≥8/1,000). **(C)** Representative network diagrams of pre-treatment intratumoural CDR3 β-chain sequences for patient ADR008. The network shows sequences that are connected to at least one other TCR within the tumour. Clustering was performed around expanded intratumoural TCRs (red circles). **(D)** The proportion of pre-treatment expanded TCRs that are part of a cluster as depicted in (C). TCRs were split between the ones that are also detected as expanded post-treatment and the ones that are not (respectively red circles and grey circles). Paired two-sided Mann–Whitney test *P* value shown. **(E**) Pre-treatment clustering around maintained and replaced expanded clones for ADR008. **(F)** The post-treatment normalised number of clusters for the networks containing expanded sequences is shown. Two-sided Mann–Whitney test *P* value shown; n=11 patients. **(G)** Representative network diagrams of post-treatment intratumoural CDR3 β-chain sequences for patient ADR001 (left) and for patient ADR013 (right). Clusters containing expanded sequences are shown.

**Figure S8. scRNA- and TCRseq of ADR013 (responder) and ADR001 (non-responder)**

**(A)** UMAP of merged ADR001 (non-responder) and ADR013 (responder) scRNA data, coloured by cell type definition (CD8 = CD8+/CD4-/FOXP3-, CD4 effector = CD8-/CD4+/FOXP3-, Treg = CD8-/FOXP3+). **(B)** Proportions of each cell type recovered in each patient. **(C)** Differential gene expression analysis performed between IgG4^+^ and IgG4^-^ cells in each cell type for each patient, average logFC then plotted for responder vs non-responder. Regression line plotted using a linear model, colours indicate whether a logFC change was found significant in either or both patients. **(D)** Heatmaps showing top genes which positively correlated (Pearson’s correlation) with TCR expansion in the non-responder (NR) patient. **(E)** Signature expression levels (calculated as the proportion of cell transcript mapping to genes in signature) by non-responder (NR) and responder (R) and IgG4 binding. Significance levels show the result of Wilcox test between IgG4 bound and unbound cells.

## Supplementary Table

**Table S1.** Baseline demographics and patient characteristics and correlations with nivolumab response, and sample annotations.

|  | All patients<br>n=15 | Responders<br>n=5 | Non-Responders<br>n=10 | p-value* |
| --- | --- | --- | --- | --- |
| **Demographics** |  |  |  |  |
| Age, median (range), years | 56 | 56 | 54 | 0.84 |
| Male, n (%) | 13 (87) | 9 (90) | 4 (80) | 0.59 |
| ECOG, n (%) |  |  |  |  |
| 0 | 8 (53) | 3 (30) | 5 (50) | 0.71 |
| 1 | 7 (70) | 2 (40) | 5 (50) |  |
| Predominant clear-cell histology, n (%) |  |  |  |  |
| Sarcomatoid/rhabdoid component, n (%) | 2 (13) | 2 (40) | 0 (0) | 0.28 |
| Prior nephrectomy, n (%) | 6 (40) | 3 (30) | 3 (60) | 0.59 |
| On-treatment nephrectomy, n(%) | 2 (13) | 1 (10) | 1 (20) | 0.59 |
| **IMDC risk categories, n (%)** |  |  |  |  |
| Favourable (0) | 2 (13) | 1 (20) | 1 (10) |  |
| Intermediate (1) | 3 (20) | 1 (20) | 2 (20) | 0.61 |
| Poor (≥2) | 10 (66) | 3 (60) | 7 (70) |  |
| **Outcomes** |  |  |  |  |
| Dead, n (%) | 6 (40) | 1 (20) | 5 (50) | 0.26 |
| Best Response by RECIST v1.1 |  |  |  |  |
| Complete response | 0 (0) | 0 (0) | 0 (0) |  |
| Partial response | 5 (33) | 4 (80) | 1 (10) | NA |
| Stable disease | 6 (40) | 1 (20) | 5 (50) |  |
| Progressive disease | 4 (27) | NA | 1 (10) |  |
| PFS, months (median) | 4.1 | 10.4 | 3.3 | 0.0007 |
| OS, months (median) | 12.5 | 12.4 | 15.0 | >0.99 |

\*Significance tests: Chi-squared test of categorical variables and Mann-Whitney U test for comparison of median values (responders vs. non-responders).
Percentages may not total 100 because of rounding.
NA - not applicable; ECOG - Eastern Cooperative Oncology Group performance scale; IMDC - International Metastatic Renal-Cell Carcinoma Database Consortium; RECIST - Response Evaluation Criteria In Solid Tumours; PFS - progression free survival; OS - overall survival

## Supplemental Data Items

**Supplemental Data Figure 1. Competition assay with anti-PD1 antibody (pembrolizumab)**. In vitro assessment of activated PBMC demonstrates that PD-1 on T cells can be detected following pembrolizumab incubation using anti-human IgG4. **(A)** Incubation of activated PBMC with pembrolizumab blocks PD-1 flow cytometry staining (EH12.2 clone). **(B)** Pembrolizumab binding to PD-1 can be detected using an anti-IgG4 flow cytometry staining antibody. All dot plots are pre-gated on live single cells.

**Supplemental Data Figure 2. Single-cell gene expression analysis of CD8**^**+**^ **and IgG4**^**+**^ **CD8**^**+**^ T-cells. Single-cell RNAseq expression of Granzyme B, TCF7, TOX, HAVCR2 (TIM-3), CD38, ENTPD1(CD39) and PDCD1(PD-1) on **(A)** CD8^+^ and (B) IgG4^+^CD8^+^T-cells in ADR013 (responder) and ADR001 (non-responder) are shown.

**Supplemental Data Table 1. Sample characteristics for: whole exome sequencing (including sequencing metrics, TMB, INDEL and neoantigen burden, purity and cancer cell fraction); RNA sequencing; TCR sequencing; and mIF/IHC**.

**Supplemental Data Table 2. Previously annotated HERV loci matched to custom repeat region annotations**. Previously annotated HERV loci from Mayer et al. and Vargui et al. and the matching loci from the custom repeat region annotation. The start and end buffer columns show the full region used for matching to the custom annotations, with 5 bases added or taken from the end or start of previously annotated loci positions respectively.

**Supplemental Data Table 3. List of genes in T-cell specific expression signatures**.

---

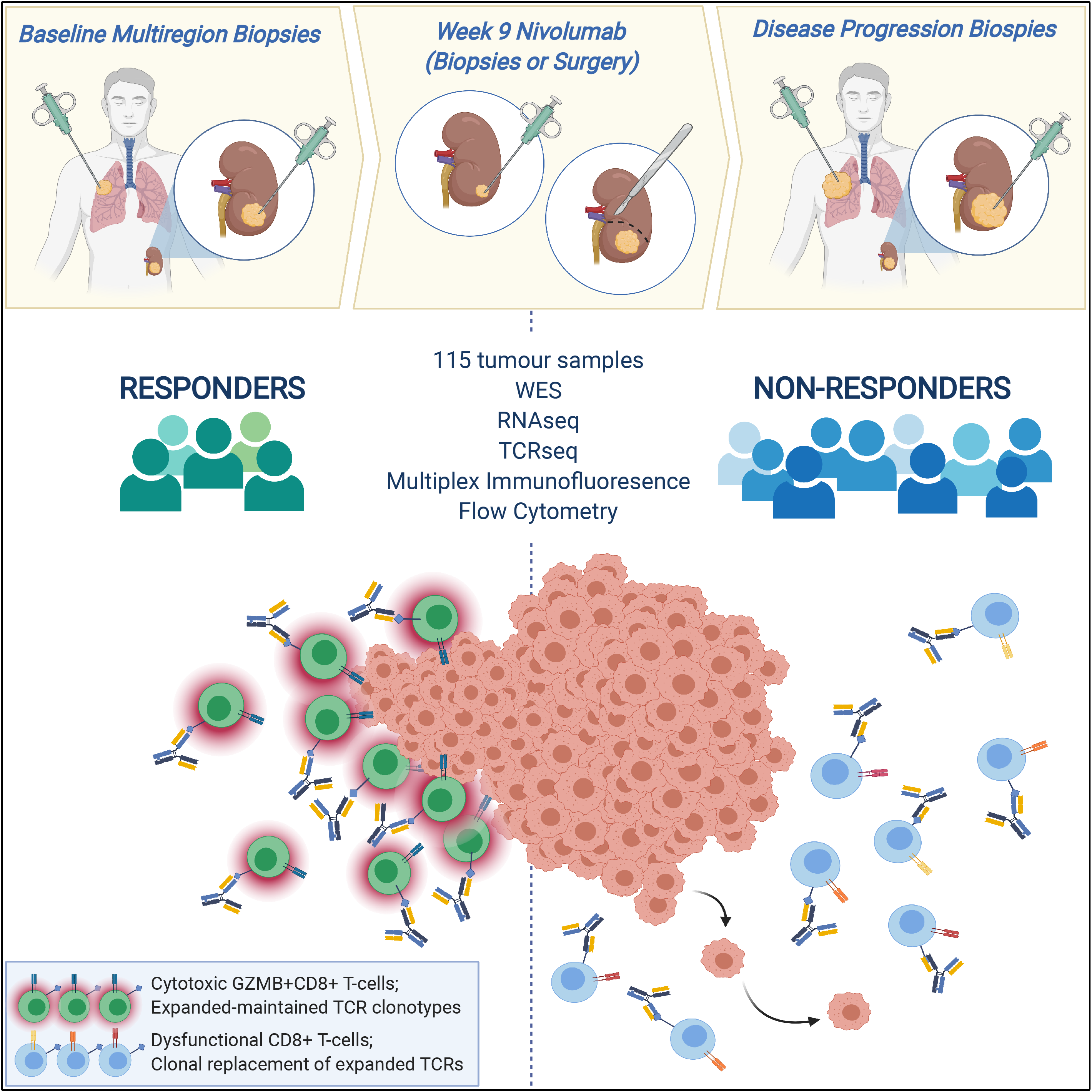

