## Supplemental Figures for "Determinants of anti-PD1 response and resistance in clear cell renal cell carcinoma"

Supplemental Material

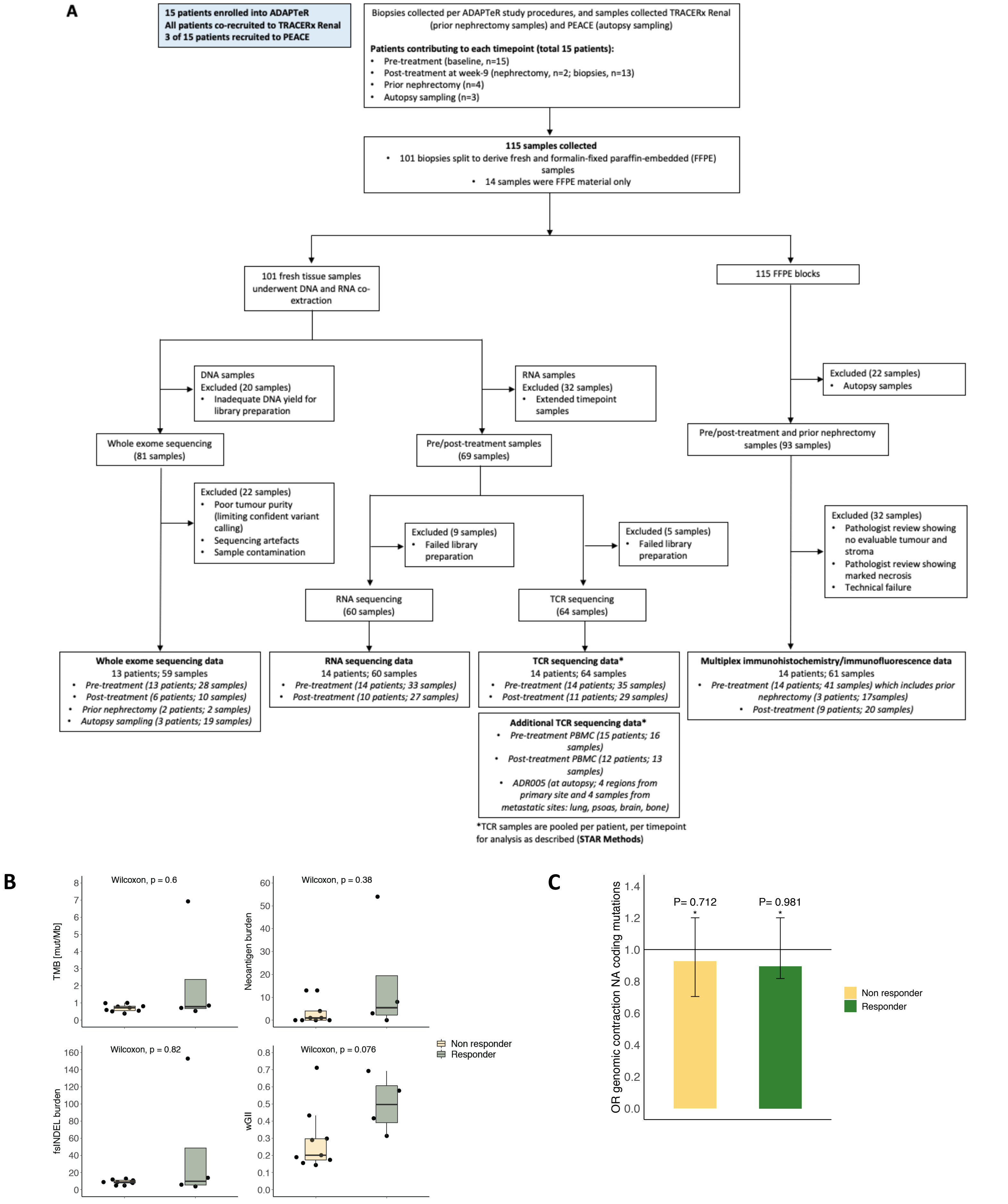


**Figure S1. Samples overview, and correlations between nivolumab response and mutational features**

**(A)** Consort diagrams for samples that underwent whole-exome sequencing, RNAseq, TCRseq, and multiplex immunofluorescence or immunohistochemistry analyses. **(B)** Boxplots showing no significant correlations between TMB, fsINDEL load, and wGII to nivolumab response. Median pre-treatment values are shown. **(C)** Genomic contraction analysis results including neoantigens. Two-sided Mann–Whitney test was performed. *P* value >0.05 considered not significant.

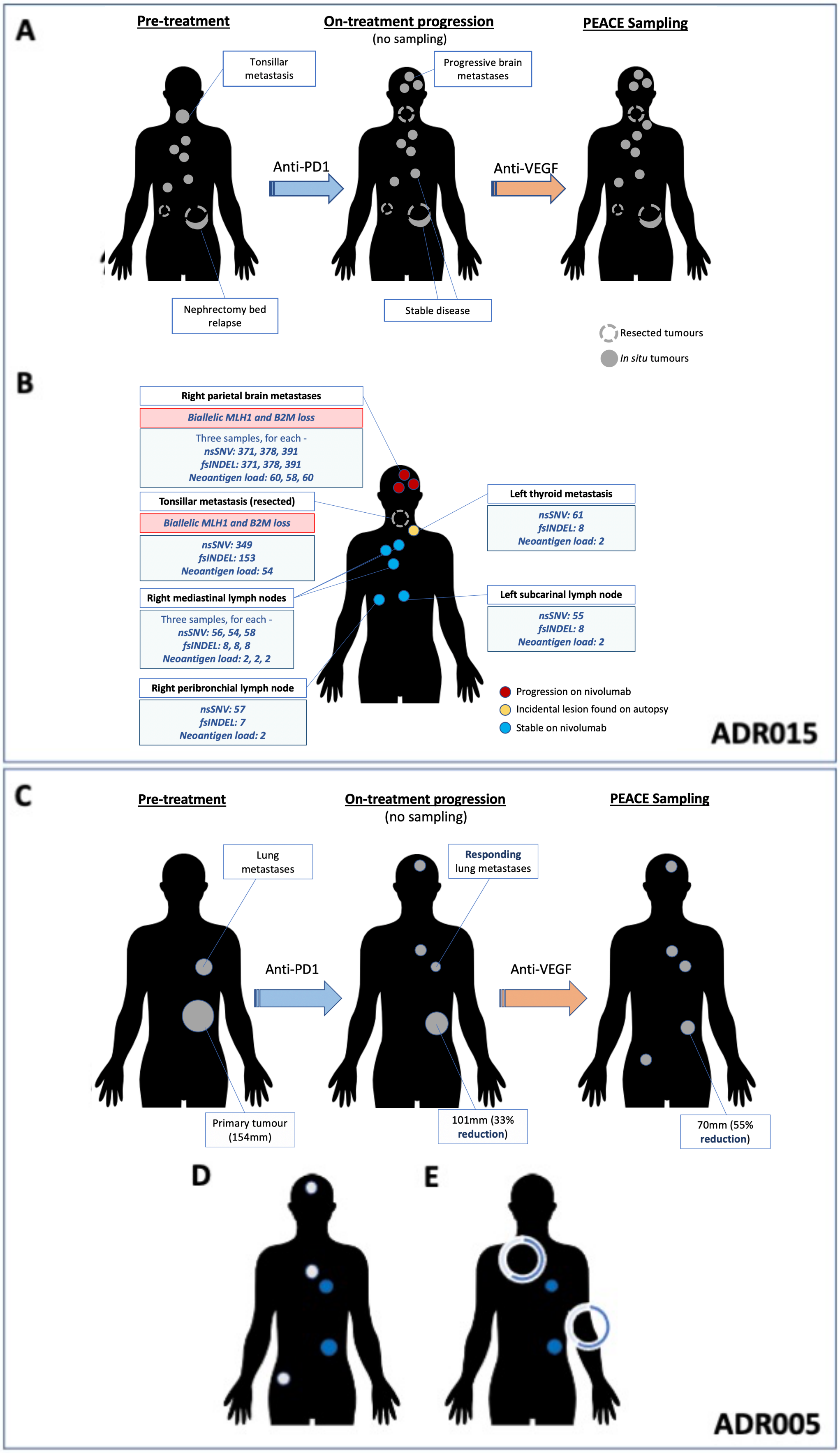


**Figure S2. Schematic diagrams of ADR015 and ADR005 showing pre-/post-treatment, post-mortem sampling, and evolution of metastatic disease** **(A)** Clinical timeline for ADR015, and **(B)** genomic data are shown. **(C)** Clinical timeline for ADR005, and **(D)** PEACE samples with available TCRseq data. **(E)** The proportion of TCRs that were expanded both Pre-treatment and Post-treatment during life (n=5) detected in each post-mortem sample, only samples where the detection rate is greater than 0 are displayed. 3/5 were detected in the lung metastatic and 1/5, 1/5, 2/5 and 3/5 were detected in region 1, region 2, region 3 and region 4 of the primary site, respectively. The median number of TCR sequences retrieved per post-mortem sample was 163 (range: 20-1340), and immune infiltration across sites were uniformly low in all regions, as scored by expert pathologist review of haematoxylin and eosin stain slides (data not shown).

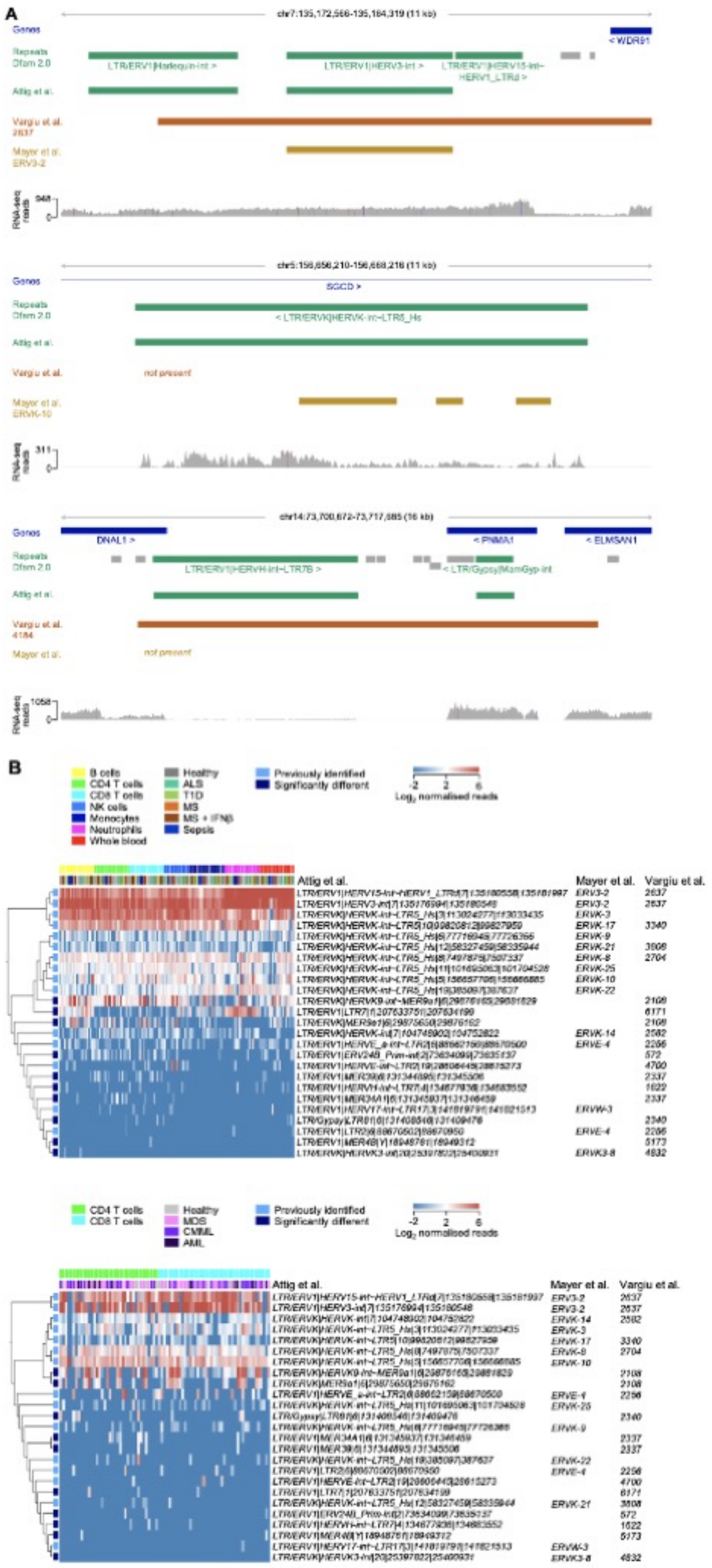

**Figure S3. Correspondence of HERV annotation and expression in immune cell types**
**(A)** Comparison of HERV annotations by Attig et al., Vargiu et al. and Mayer et al. Three examples are shown, depicting the position of GENCODE annotated genes, Dfam 2.0 annotated repeats and a representative RNAseq read pileup. **(B)** Expression of HERVs previously associated with ccRCC or with nivolumab response, in the indicated purified immune cell types from public RNAseq datasets GSE60424 (top) and E-MTAB-8208 (bottom). Note the expression of LTR/ERVK|HERVK9-int~MER9a1|6|29876165|29881829 integration within the HLA locus in most immune cell subsets and of the LTR/ERV1|LTR7|1|207633751|207634199 integration in neutrophils.

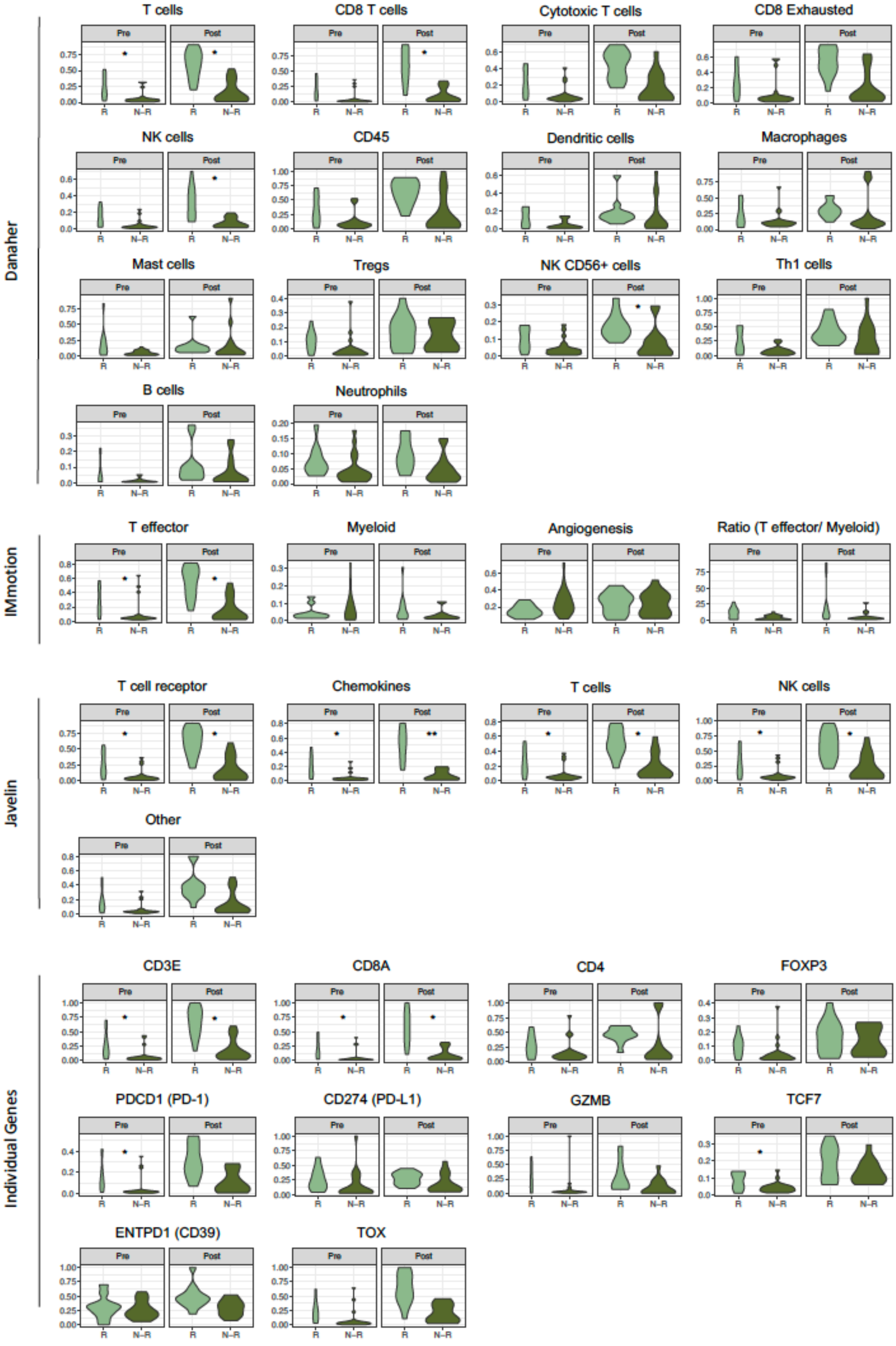


**Figure S4. Violin plots comparing response groups at both timepoints by Danaher, IMMOTION150, Javelin101 signatures and by individual gene expression.**

See **STAR Methods** for details of signature analysis. The two-sided Mann–Whitney test performed on one value per patient (score averaged by median value across biopsies if several available at a given time point), significant *P* value are indicated (*: *P*<0.05; **: *P*<0.01). *R - responders; N-R - non-responders.*

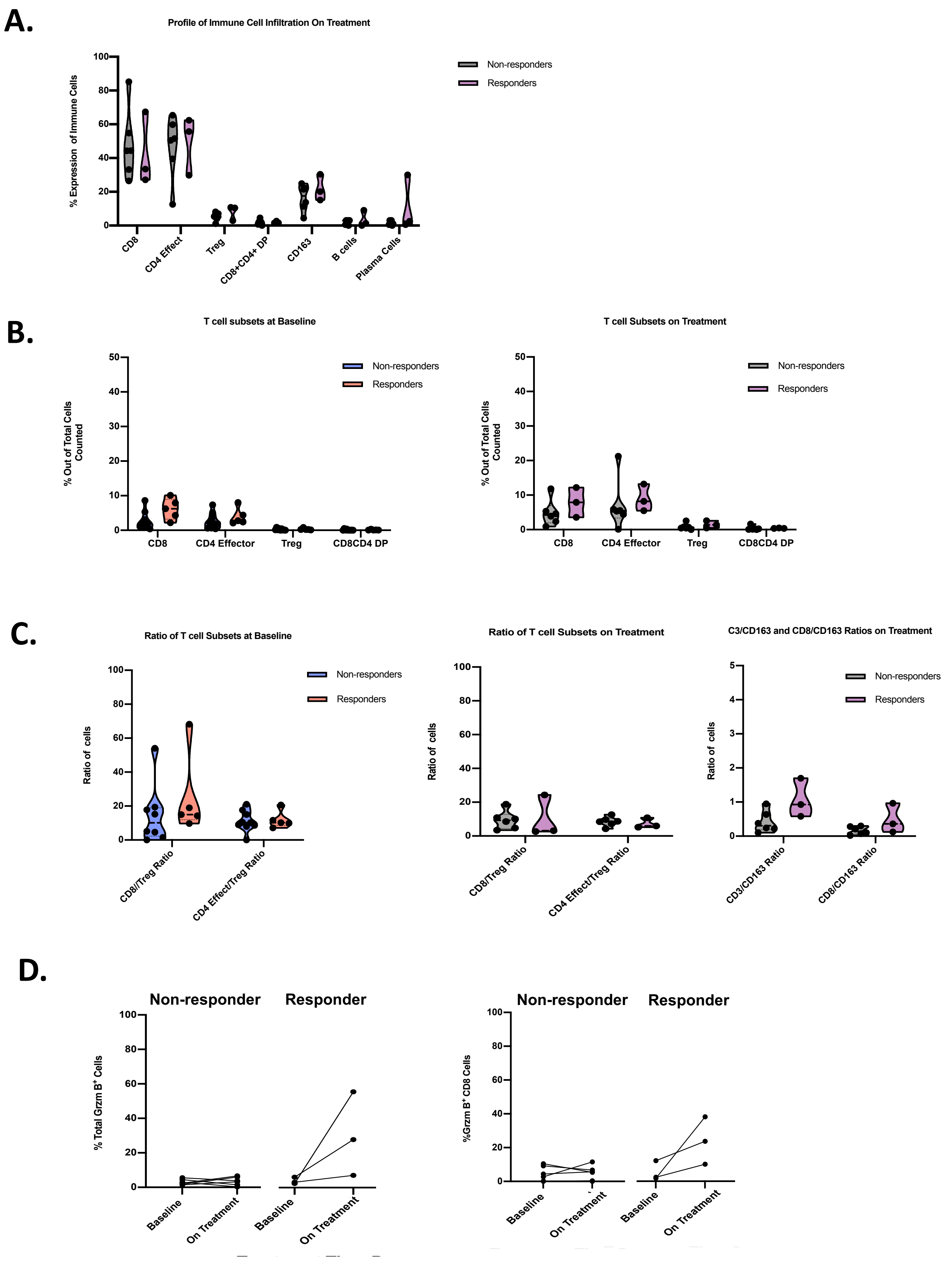


**Figure S5. Immune cells subset comparisons of pre- and post-treatment samples**

**(A)** Immune cell subset expression levels in non-responders and responders on treatment are shown. **(B)** Expression level of T-cell subsets out of total cells counted is shown. **(C)** Ratio of T-cells subsets in non-responders and responders at baseline and on treatment; $CD3^+$ T-cells to $CD163^+$ myeloid cells, and $CD8^+$ T-cells to $CD163^+$ myeloid cell ratios between responders and non-responders on treatment is shown. **(D)** Change in total GZMB expression and on $CD8^+$ T cells from pre-treatment (six patients) to post-treatment (week-9) is shown (three patients).

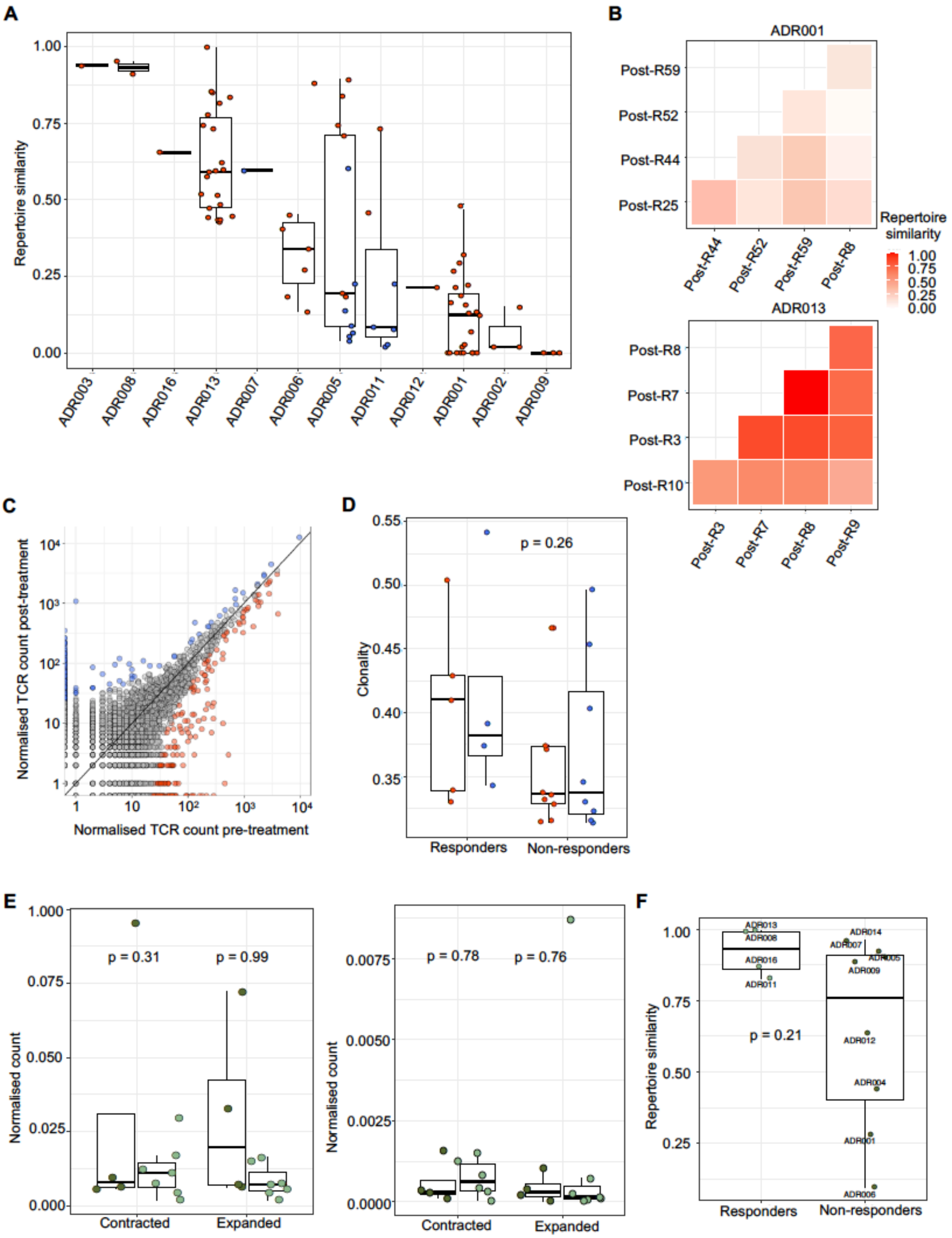


**Figure S6. Clonotype dynamics in PBMC and intra- and inter-patient TCR repertoire heterogeneity**

**(A)** The TCR repertoires of multiple biopsies from a patient's tumor were sequenced and a pairwise comparison of the repertoires of different biopsies from the same timepoint was performed by using the cosine metric (**STAR Methods**). The pairwise intratumoral TCR repertoire similarity is shown for each patient. Each circle represents a comparison between two samples from the same patient (*n* = 87 total comparisons from 12 patients). Red (resp. blue) circles indicate a pair of biopsies originating from the same site (resp. different metastatic sites). **(B)** Heat maps showing the pairwise similarities of a selection of 5 biopsies in the post-treatment nephrectomy for ADR001 (top) and ADR013 (bottom). Biopsies were selected based on comparable TCR counts. **(C)** Correlated clone sizes in blood samples. Scatter plots of blood clone size after treatment and before treatment are shown for all patients. Clones are colored by expansion/contraction status (**STAR Methods**). **(D)** The peripheral TCR repertoire clonality score pre-treatment and on-treatment is shown for each patient. Patients are split between responders and non-responders. Mixed-effect model *P* value shown. **(E)** The number of intratumoral (left panel) and peripheral (right panel) clones labelled as expanded or contracted between timepoints, per patient, normalized for the total number of clones tested. Two-sided Mann–Whitney tests *P* value shown. **(F)** The peripheral cosine score between pre-treatment and on-treatment is shown for each patient. Patients are split between responders and non-responders. Two-sided Mann–Whitney test *P* value shown; n=12 patients.

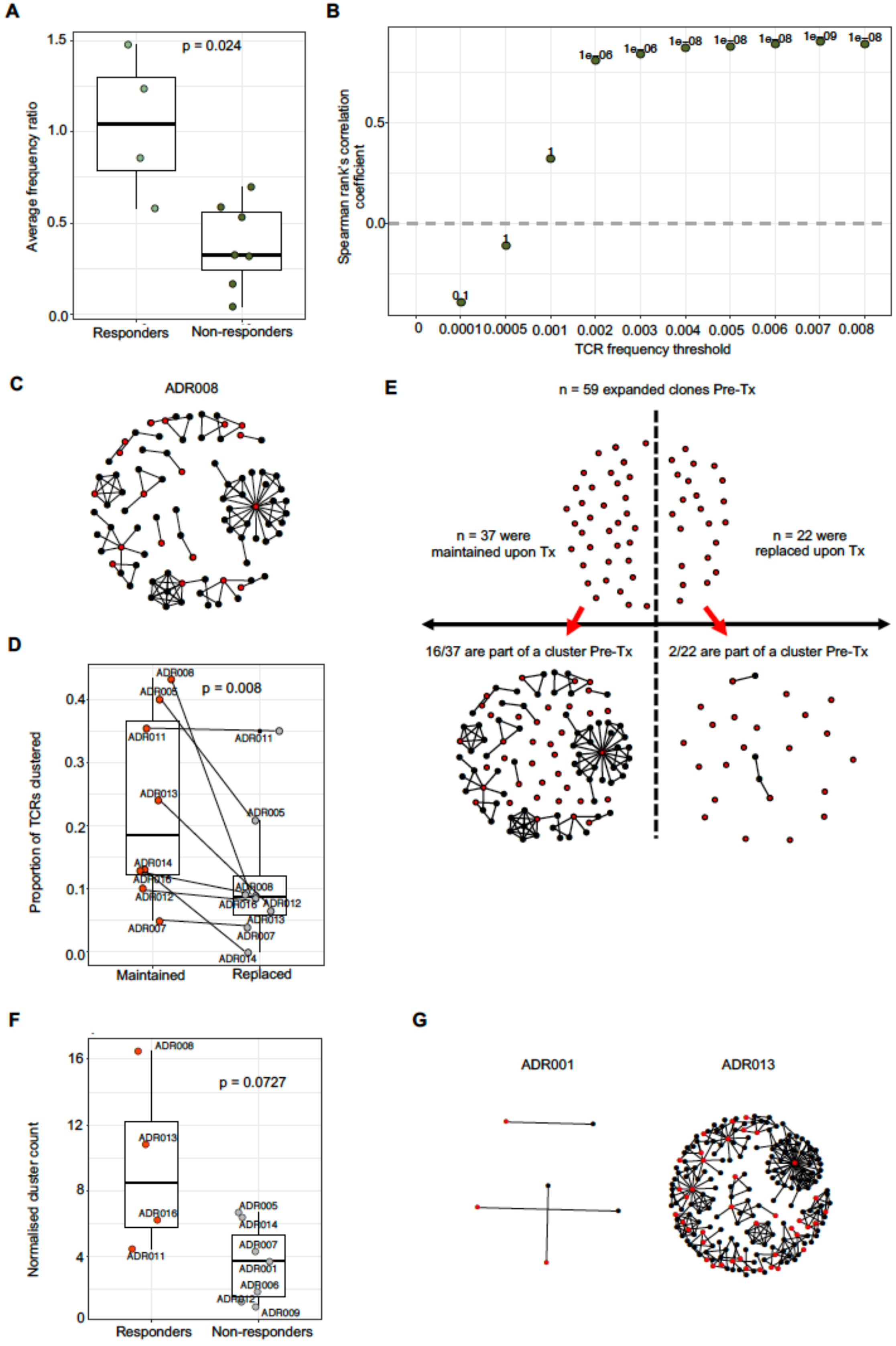


**Figure S7. Additional expanded TCRs metrics**

**(A)** The arithmetic mean of Pre/Post frequency ratios of clones expanded pre-treatment, per patient. Two-sided Mann–Whitney test *P* value shown. **(B)** The Spearman's rank correlation coefficient and *P* value (shown above each point; n=14 patients) for the relationship between the clonality score and the proportion of the intratumoral repertoire pre-treatment occupied by expanded clones defined by different frequency thresholds (ranging from all TCRs (threshold of zero) up to those found at a frequency of ≥8/1,000). **(C)** Representative network diagrams of pre-treatment intratumoral CDR3 β-chain sequences for patient ADR008. The network shows sequences that are connected to at least one other TCR within the tumor. Clustering was performed around expanded intratumoral TCRs (red circles). **(D)** The proportion of pre-treatment expanded TCRs that are part of a cluster as depicted in (C). TCRs were split between the ones that are also detected as expanded post-treatment and the ones that are not (respectively red circles and grey circles). Paired two-sided Mann–Whitney test *P* value shown. **(E)** Pre-treatment clustering around maintained and replaced expanded clones for ADR008. **(F)** The post-treatment normalised number of clusters for the networks containing expanded sequences is shown. Two-sided Mann–Whitney test *P* value shown; n=11 patients. **(G)** Representative network diagrams of post-treatment intratumoral CDR3 β-chain sequences for patient ADR001 (left) and for patient ADR013 (right). Clusters containing expanded sequences are shown.

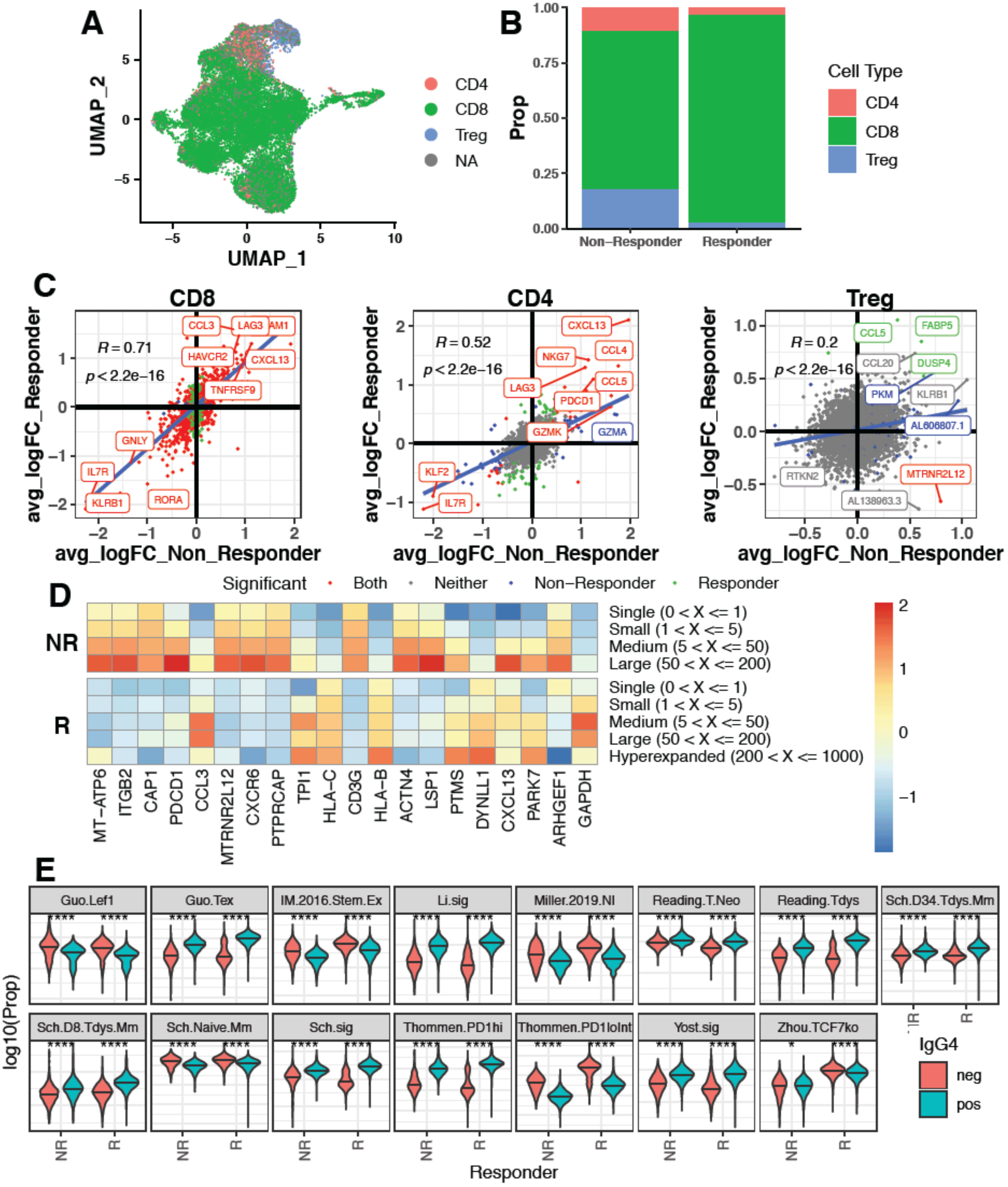

**Figure S8. scRNA- and TCRseq of ADR013 (responder) and ADR001 (non-responder)**

**(A)** UMAP of merged ADR001 (non-responder) and ADR013 (responder) scRNA data, coloured by cell type definition (CD8 = CD8+/CD4-/FOXP3-, CD4 effector = CD8-/CD4+/FOXP3-, Treg = CD8-/FOXP3+). **(B)** Proportions of each cell type recovered in each patient. **(C)** Differential gene expression analysis performed between IgG4$^{+}$ and IgG4$^{-}$ cells in each cell type for each patient, average logFC then plotted for responder vs non-responder. Regression line plotted using a linear model, colours indicate whether a logFC change was found significant in either or both patients. **(D)** Heatmaps showing top genes which positively correlated (Pearson's correlation) with TCR expansion in the non-responder (NR) patient. **(E)** Signature expression levels (calculated as the proportion of cell transcript mapping to genes in signature) by non-responder (NR) and responder (R) and IgG4 binding. Significance levels show the result of Wilcox test between IgG4 bound and unbound cells.

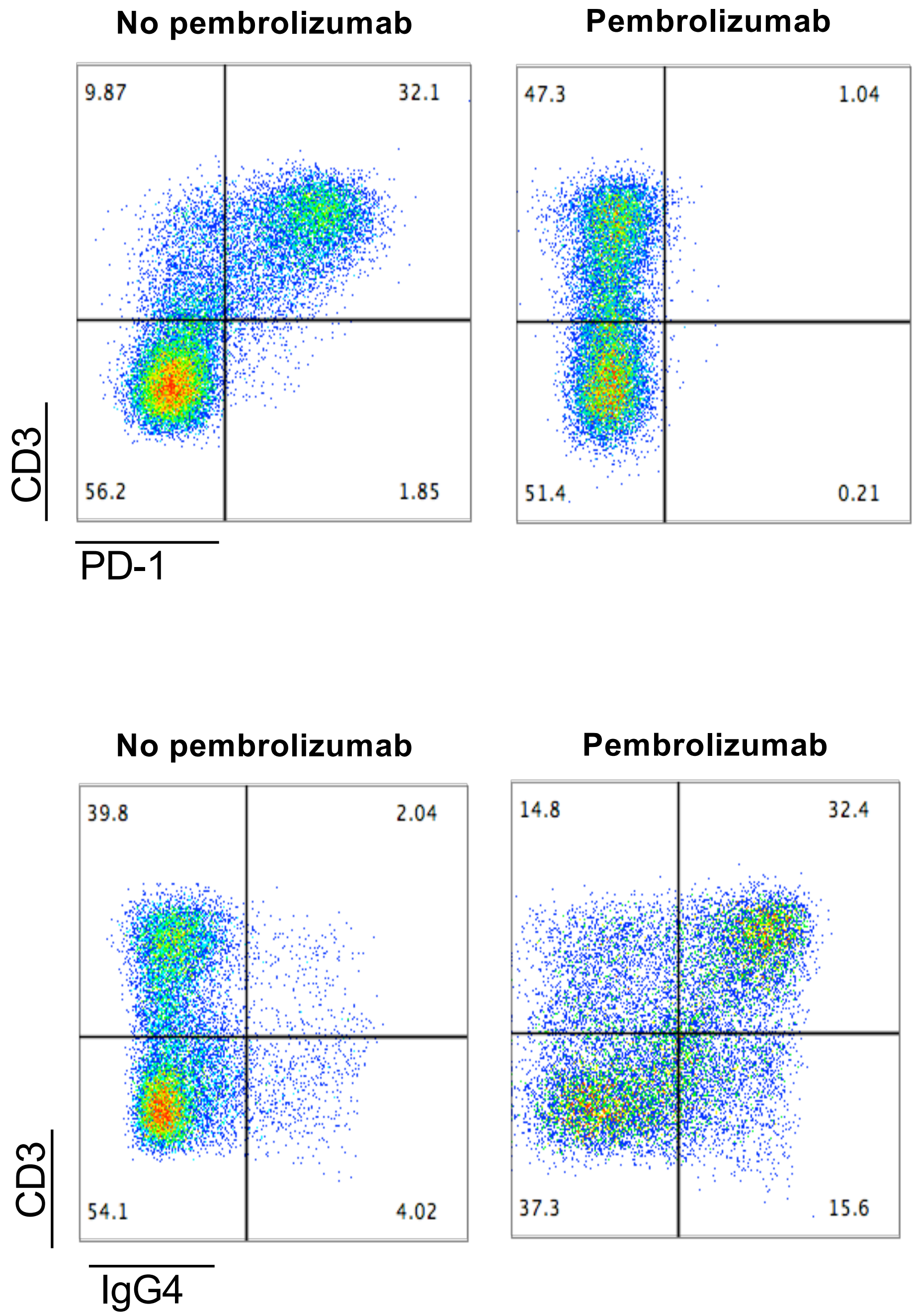


**Supplemental Data Figure 1. Competition assay with anti-PD1 antibody (pembrolizumab).** In vitro assessment of activated PBMC demonstrates that PD-1 on T cells can be detected following pembrolizumab incubation using anti-human IgG4. **(A)** Incubation of activated PBMC with pembrolizumab blocks PD-1 flow cytometry staining (EH12.2 clone). **(B)** Pembrolizumab binding to PD-1 can be detected using an anti-IgG4 flow cytometry staining antibody. All dot plots are pre-gated on live single cells.

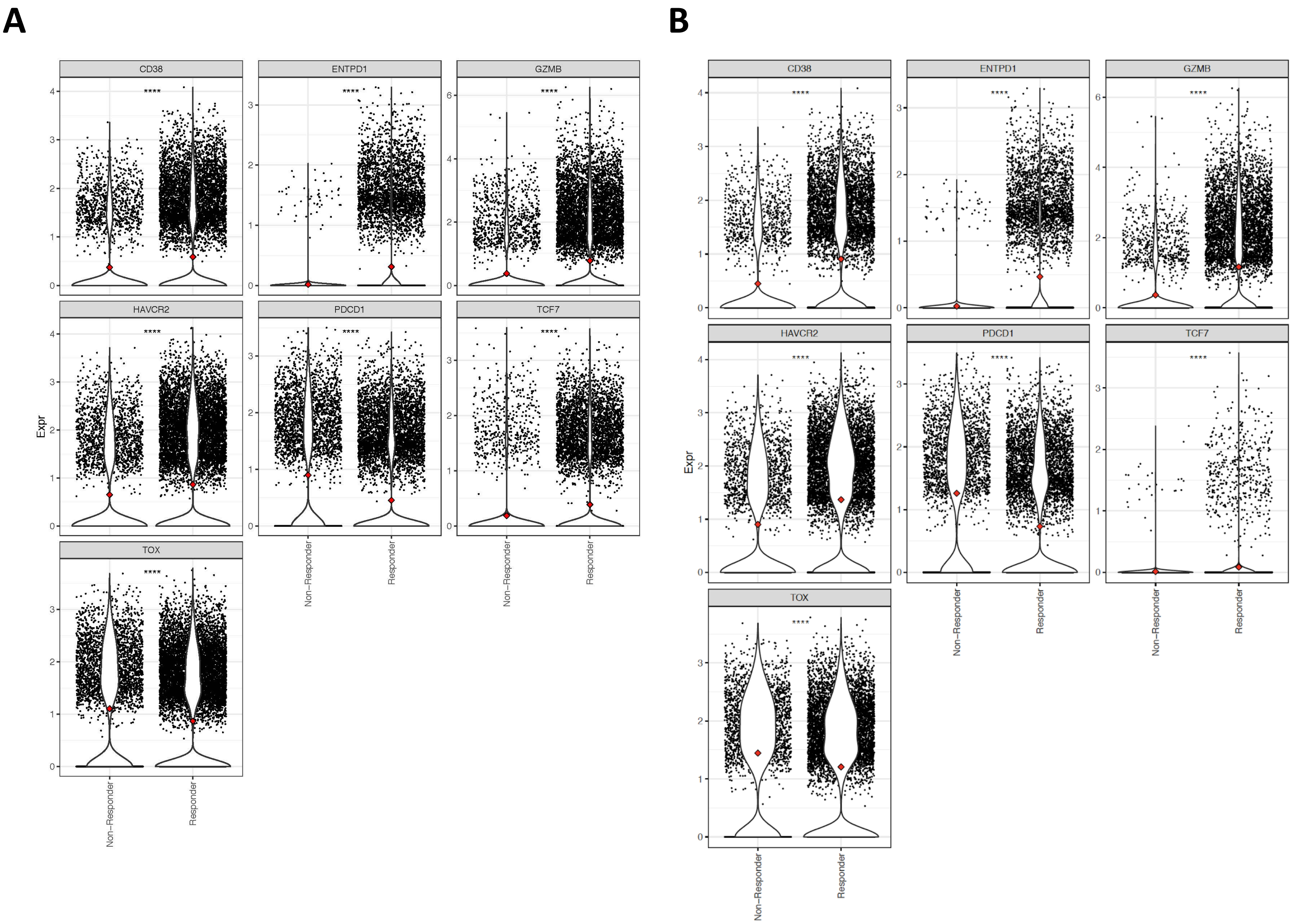


**Supplemental Data Figure 2. Single-cell gene expression analysis of CD8⁺ and IgG4⁺CD8⁺ T-cells.** Single-cell RNAseq expression of Granzyme B, TCF7, TOX, HAVCR2 (TIM-3), CD38, ENTPD1(CD39) and PDCD1(PD-1) on **(A)** $CD8^+$ and **(B)** $IgG4^+CD8^+$ T-cells in ADR013 (responder) and ADR001 (non-responder) are shown.
